# A literature scanning and prioritization framework to guide future systematic reviews for World Cancer Research Fund International’s Global Cancer Update Programme

**DOI:** 10.64898/2026.05.07.26352530

**Authors:** Georgios Markozannes, Ahmad Jayedi, Sofia Cividini, Sayada Zartasha Kazmi, Margarita Cariolou, Rita Vieira, Eirini Pagkalidou, Sonia Kiss, Katia Balducci, Dagfinn Aune, Marc J. Gunter, Amanda J. Cross, Doris S. M. Chan, Konstantinos K. Tsilidis

## Abstract

**Background:** The 2018 World Cancer Research Fund (WCRF)/American Institute for Cancer Research Third Expert Report (TER) on diet, adiposity, physical activity and risk of 19 cancers could be enhanced with new data. A framework is needed to prioritize future systematic reviews.

**Methods:** We searched PubMed (January 2019-February 2024) for meta-analyses, pooled analyses, randomized controlled trials (RCTs), Mendelian randomization (MR) studies, and large (>100,000 participants) cohort studies. We assessed TER findings using conditional power (CP) and fail-safe number (FSN) statistics. We developed an exposure-based prioritization score (PS) by awarding or subtracting points considering the quantity, statistical significance, direction, and novelty of associations.

**Results:** We compared 366 meta-analyses, 121 pooled analyses, 19 RCTs, 174 MR studies, and 391 cohort studies covering 151 exposures and 28 cancers with 1,371 TER meta-analyses. Based on CP, non-significant TER associations likely to become significant with additional evidence included folate and colorectal, waist circumference and lung, total fat and ovarian, tea and ovarian, and red meat and kidney cancers. The FSN indicated that most significant TER associations are unlikely to change with additional evidence. The median PS was 6 (range: −15 to 163), with top scores observed for anthropometric measurements (PS_height_=40 to PS_BMI_=163), physical activity (PS=100), sedentary behavior (PS=64), alcohol (PS=52), tea (PS=36), dietary fiber (PS=31), milk/dairy (PS=29), micronutrients (PS_retinol_=27 to PS_iron_=38), vitamins (PS_B12_=22 to PS_vitD_=91), soy (PS=24), isoflavones (PS=23), and sugar-sweetened beverages (PS=22).

**Conclusions and Impact:** The prioritization framework can help identify impactful systematic reviews to complement TER conclusions and enhance our understanding of emerging research.

## Introduction

Cancer remains a leading cause of morbidity and mortality globally, with an estimated 20 million new cases and 9.7 million cancer deaths in 2022 **(1)**. Since 1997, the World Cancer Research Fund (WCRF) network have synthesized scientific evidence on modifiable lifestyle factors, such as adiposity, diet, physical activity, and sedentary behavior in relation to cancer risk, contributing to evidence-based recommendations for cancer prevention **(2)**. The Global Cancer Update Programme (CUP Global) is WCRF International’s flagship programme that systematically reviews global research to support the recommendations. An independent Expert Panel grades the CUP Global evidence using predefined criteria evaluating the likelihood of causality as strong (convincing; probable; substantial effect on risk unlikely) or limited (suggestive; no conclusion). Strong evidence typically forms the basis for cancer prevention recommendations. The Third Expert Report (TER) **(3)** published in 2018 utilized evidence from cohort studies and randomized controlled trials (RCTs) to provide a comprehensive assessment of the role of lifestyle factors across 19 cancers. Over 700 exposure-cancer associations were examined and 23 were classified with convincing, 37 probable, and 64 limited-suggestive evidence. This evidence was used to develop the 2018 WCRF/American Institute for Cancer Research (AICR) cancer prevention recommendations **(4)**, which have shaped global health policies and guided individuals in reducing their cancer risk.

Comprehensive, timely, and up-to-date evidence assessments are crucial for effective preventive strategies and public health policies to reduce cancer burden. Many associations in the TER were graded with limited-no conclusion evidence, largely due to sparse data. Since the publication of the TER, a considerable number of relevant meta-analyses and pooled analyses have been published. This accumulation of data provides opportunities to assess emerging research directions, such as life-course exposures, dietary and lifestyle patterns, and cancer subtypes, while also revising past conclusions and clarifying previous less-explored associations **(1)**. Furthermore, there are now data on less well studied cancers, such as brain cancer, thyroid cancer, lymphomas, myelomas and leukemias **(5–8)**. In recent years, studies from Asian countries have contributed valuable findings **(9–20)**, partly addressing the research gap related to studies of non-white ethnicity.

CUP Global’s systematic literature reviews (SLRs) are comprehensive but resource-intensive and time-consuming, necessitating more efficient strategies to prioritize future SLR topics. To address this, we developed a prioritization framework to support the identification and ranking of SLRs that have accumulated sufficient evidence to complement the TER conclusions and enhance our understanding of emerging topics. This paper provides an overview of the accumulated literature, along with a framework designed to identify, rank and prioritize research areas for new and updated SLRs, providing the basis for a structured five-year plan for CUP Global’s SLRs.

## Methods

The methodology below outlines the actions that were utilized for identifying and prioritizing research topics on the association between adiposity, diet, physical activity, sedentary behavior and cancer risk. Rather than surveying all new studies, we targeted specific research designs to identify trends, evidence gaps, and priority areas for future SLRs. This framework involved six actions: scanning research published since the TER, statistically assessing the robustness of TER findings, and creating a scoring algorithm to rank and prioritize research topics for future SLRs (**Figure 1**). The scanning included:

**Figure 1:**
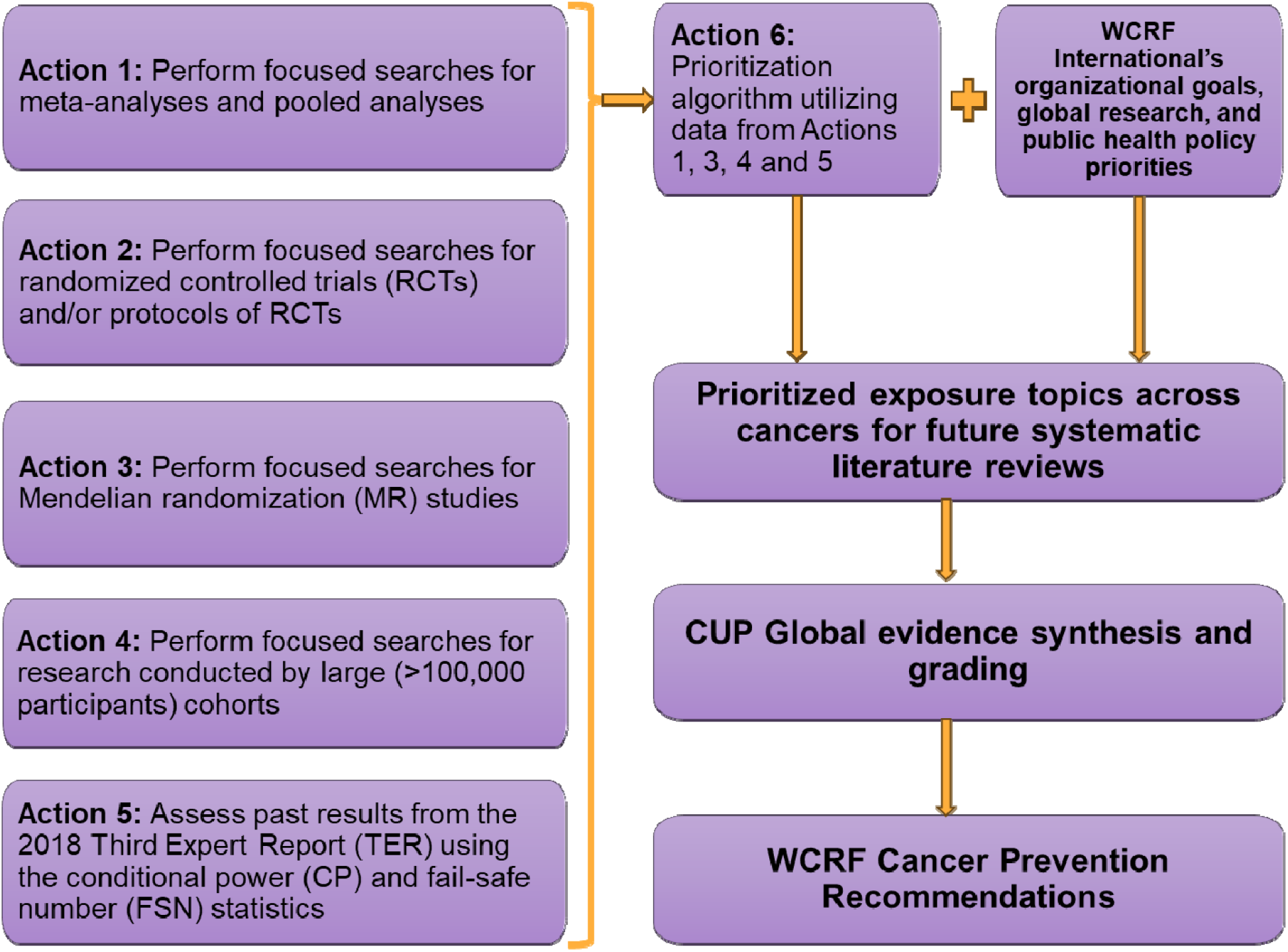
Overview of the CUP Global data scanning and prioritization framework ^a^. Abbreviations: CUP Global: Global Cancer Update Programme, WCRF: World Cancer Research Fund. ^a^ Results from RCTs were not included in the prioritization algorithm, as most were classified as mixed interventions, could not be aligned to a specific exposure or cancer site, or were not statistically significant.

### Action 1: Focused searches for systematic reviews, meta-analyses and pooled analyses

This action identifies previously covered topics with accumulated literature and highlights opportunities to revisit topics from the TER, providing insights for novel or updated SLRs.

### Action 2: Focused searches for RCTs and/or protocols of RCTs

Although rare, RCTs with cancer incidence as the primary endpoint are critical within CUP Global’s grading scheme, potentially impacting future evidence assessments. Similarly, trials on cancer precursors (e.g., adenomatous polyps, serrated polyps, cervical intraepithelial neoplasia) are also relevant. Identifying RCT protocols and evaluating their completion dates can aid future SLR planning.

### Action 3: Focused searches for Mendelian randomization (MR) studies

MR studies have a complementary role in triangulating findings from other actions and offer insights on mechanisms and potential novel associations.

### Action 4: Focused searches for large cohort studies

Results from large cohort studies often guide meta-research directions. We focused on thirteen large-scale (over 100,000 participants) cohorts from the Pooling Project of Prospective Studies of Diet and Cancer and major biobanks exploring lifestyle factors and cancer risk (**Supplementary Text I**).

### Search strategy and study selection (actions 1-4)

For actions 1 to 4, we searched PubMed (January 2019 to February 2024) using tailored search algorithms based on CUP Global search strategies (**Supplementary Text II**). This period was selected to capture recent and emerging evidence following the TER, focusing on studies likely to inform future SLR priorities rather than exhaustively identifying all publications. Screening and study selection was performed by a single reviewer. An independent reviewer assessed at least 10% of the identified studies for accuracy.

### Data extraction (actions 1-4)

We recorded all exposure-cancer associations and captured basic data for each association, including the exposure of interest (allocated into major exposure groups), cancer site (or subsite/subtype), outcome type (incidence/mortality among healthy populations), association type (dose-response, categorical), subgroup analyses (e.g., by sex, ethnicity, smoking status), direction and statistical significance of the association. For action 1 (meta-analyses, pooled analyses), we also captured search dates, study designs (cohort, case-control), and whether a meta-analysis had been performed. For action 2 (RCTs), we further recorded trial names, design, blinding, interventions, and from protocols expected completion dates and publication plans. In action 3 (MR studies), we additionally captured population ancestries and instrumental variable details. For action 4 (cohorts), we recorded the study names. Data extraction was performed by one reviewer and verified by a second independent reviewer.

### Action 5: Statistical assessment of the TER findings

We statistically assessed the TER findings to determine whether future research could alter the TER conclusions. For non-statistically significant meta-analytic associations, we computed the conditional power (CP) to assess if future studies similar to those in the TER meta-analyses may affect inference **(21)**. A very high estimated number of future studies (based on 80% CP) in relation to the number of studies in the TER meta-analysis indicates that further research might be futile or the need for considerably larger studies, while a low number suggests that even limited additional research could affect the TER conclusions. For statistically significant meta-analytic associations, we computed the fail-safe number (FSN), representing the number of additional “null” studies needed to nullify the observed meta-analytic estimate **(22)**. A high FSN indicates possible stability of findings, while a low FSN indicates that inference could change with only a few additional null studies.

### Acton 6: Data prioritization framework

We developed a structured framework to prioritize future SLRs by ranking research topics based on the quantity, statistical significance, and direction of the new study estimates. By assigning “prioritization” points to each exposure-cancer association, we created a prioritization score (PS) to facilitate strategic decision-making in liaison with the WCRF International’s CUP Global Secretariat about which areas to prioritize for future SLRs.Topics with the potential to upgrade evidence conclusions from the TER or those introducing new, previously unexamined information had higher PS and were prioritized for discussions with WCRF International about future reviews, also considering organizational and global research and public health policy priorities.

Details for the rationale and methodology of all actions are provided below and expanded in the **Supplement**.

The prioritization algorithm is summarized in **Table 1**. Detailed description and rationale are provided in **Supplementary Text III**. Briefly, the algorithm compared the results from actions 1, 3, and 4 with the TER findings (action 5), prioritizing exposure-cancer associations that were concordant in direction (regardless of magnitude) and in statistical significance with those of the TER. We did not incorporate RCTs in the PS, as most RCTs either investigated mixed interventions or cancer sites that could not be directly aligned to associations of interest or reported non-significant associations (possibly due to lack of power). Associations showing concordant direction with the TER received points, while those with discordant direction were subtracted points, assuming that the addition of this data was unlikely to strengthen already established conclusions. Associations were awarded or subtracted points from CP analyses, depending on whether the observed number of studies in the new meta-analyses (action 1) and in the TER (action 5) exceeded or not the estimated number required to change the TER results. While the FSN was initially considered for the scoring algorithm, it was ultimately not used, as it proved overly conservative and would have penalized smaller meta-analyses with lower evidence grading on important topics that could benefit from additional research despite meaningful and consistent accumulation of new evidence. Instead, it was used as an additional analysis to ensure completeness. Non-significant results from MR analyses were not used to subtract points due to potential statistical power issues. Novel associations (e.g., for new cancer sites or sub-analyses) not investigated in the TER that could support a meta-analysis with at least three independent studies were also awarded points. Within the CUP Global SLRs, exposures such as dietary and lifestyle patterns are often descriptively synthesized without a meta-analysis, due to their inherent heterogeneity in terms of their individual components. These exposures, while being included in the prioritization framework, were excluded from the PS calculations.

**Table 1:**
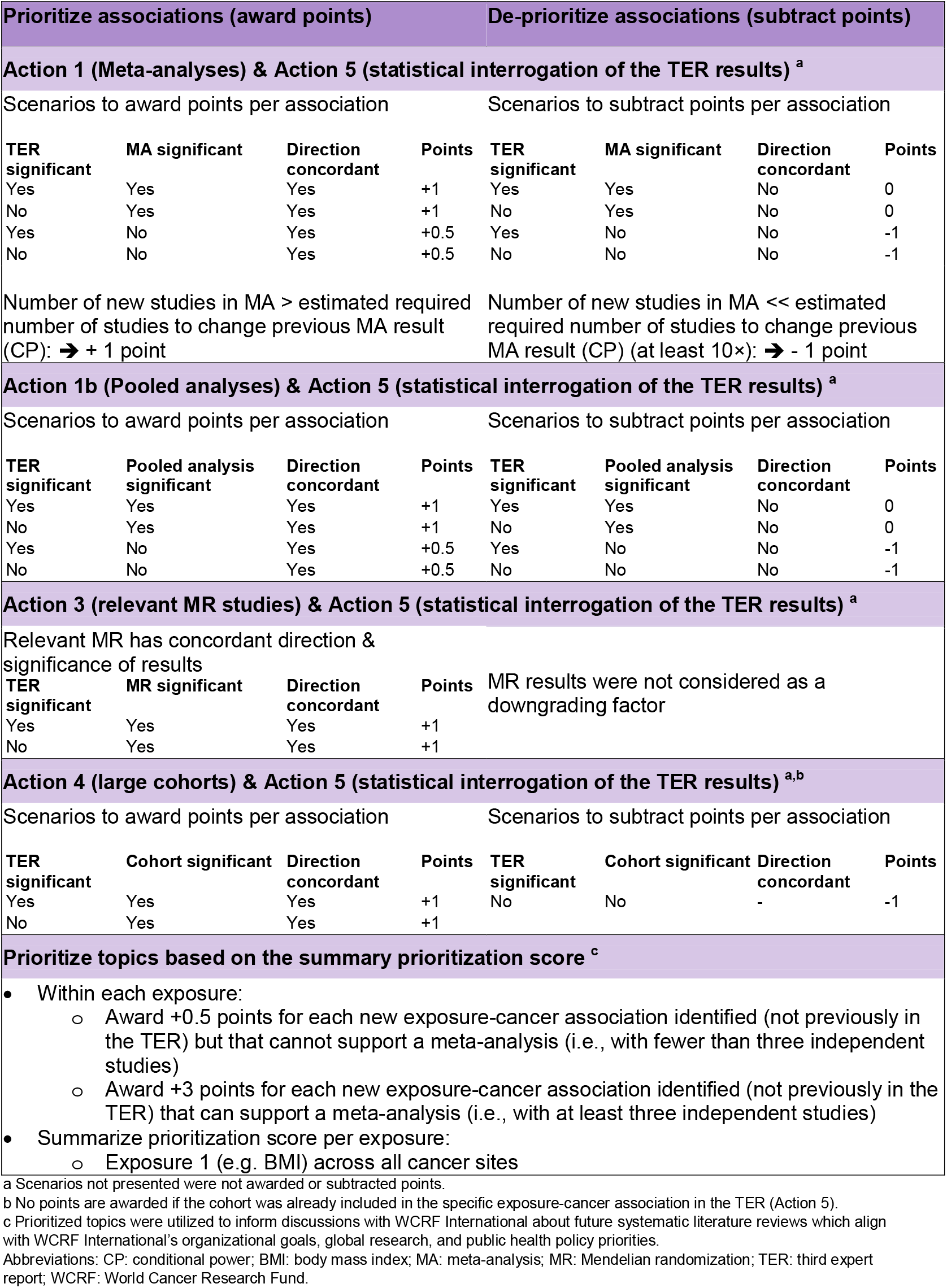
Summary of the prioritization algorithm.

| Prioritize associations (award points) |  |  |  | De-prioritize associations (subtract points) |  |  |  |
| --- | --- | --- | --- | --- | --- | --- | --- |
| Action 1 (Meta-analyses) & Action 5 (statistical interrogation of the TER results)<sup>a</sup> |  |  |  |  |  |  |  |
| Scenarios to award points per association |  |  |  | Scenarios to subtract points per association |  |  |  |
| TER significant | MA significant | Direction concordant | Points | TER significant | MA significant | Direction concordant | Points |
| Yes | Yes | Yes | +1 | Yes | Yes | No | 0 |
| No | Yes | Yes | +1 | No | Yes | No | 0 |
| Yes | No | Yes | +0.5 | Yes | No | No | -1 |
| No | No | Yes | +0.5 | No | No | No | -1 |
| Number of new studies in MA > estimated required number of studies to change previous MA result (CP): ➔ + 1 point |  |  |  | Number of new studies in MA << estimated required number of studies to change previous MA result (CP) (at least 10×): ➔ - 1 point |  |  |  |
| Action 1b (Pooled analyses) & Action 5 (statistical interrogation of the TER results)<sup>a</sup> |  |  |  |  |  |  |  |
| Scenarios to award points per association |  |  |  | Scenarios to subtract points per association |  |  |  |
| TER significant | Pooled analysis significant | Direction concordant | Points | TER significant | Pooled analysis significant | Direction concordant | Points |
| Yes | Yes | Yes | +1 | Yes | Yes | No | 0 |
| No | Yes | Yes | +1 | No | Yes | No | 0 |
| Yes | No | Yes | +0.5 | Yes | No | No | -1 |
| No | No | Yes | +0.5 | No | No | No | -1 |
| Action 3 (relevant MR studies) & Action 5 (statistical interrogation of the TER results)<sup>a</sup> |  |  |  |  |  |  |  |
| Relevant MR has concordant direction & significance of results |  |  |  | MR results were not considered as a downgrading factor |  |  |  |
| TER significant | MR significant | Direction concordant | Points |  |  |  |  |
| Yes | Yes | Yes | +1 |  |  |  |  |
| No | Yes | Yes | +1 |  |  |  |  |
| Action 4 (large cohorts) & Action 5 (statistical interrogation of the TER results)<sup>a,b</sup> |  |  |  |  |  |  |  |
| Scenarios to award points per association |  |  |  | Scenarios to subtract points per association |  |  |  |
| TER significant | Cohort significant | Direction concordant | Points | TER significant | Cohort significant | Direction concordant | Points |
| Yes | Yes | Yes | +1 | No | No | - | -1 |
| No | Yes | Yes | +1 |  |  |  |  |
| Prioritize topics based on the summary prioritization score<sup>c</sup> |  |  |  |  |  |  |  |
| Within each exposure:<br>Award +0.5 points for each new exposure-cancer association identified (not previously in the TER) but that cannot support a meta-analysis (i.e., with fewer than three independent studies)<br>Award +3 points for each new exposure-cancer association identified (not previously in the TER) that can support a meta-analysis (i.e., with at least three independent studies)<br>Summarize prioritization score per exposure:<br>Exposure 1 (e.g. BMI) across all cancer sites |  |  |  |  |  |  |  |

<sup>a</sup> Scenarios not presented were not awarded or subtracted points.
<sup>b</sup> No points are awarded if the cohort was already included in the specific exposure-cancer association in the TER (Action 5).
<sup>c</sup> Prioritized topics were utilized to inform discussions with WCRF International about future systematic literature reviews which align with WCRF International's organizational goals, global research, and public health policy priorities.
Abbreviations: CP: conditional power; BMI: body mass index; MA: meta-analysis; MR: Mendelian randomization; TER: third expert report; WCRF: World Cancer Research Fund.

A PS was calculated at the exposure-cancer level by summing scores across all actions while also incorporating points from new cancer sites, subgroups, or populations. These scores were further summed to create a total exposure-specific PS representing the overall alignment and support for each association across actions with the TER findings as the reference point. Exposures were then presented by decreasing PS (overall and by cancer site within exposures) indicating higher to lower priorities for potential future SLRs.

As sensitivity analyses, we excluded associations previously graded as “strong-convincing” in the TER, assuming these topics require less immediate attention, and also calculated cancer-specific PS by aggregating scores across all exposures. For novel exposures that were not previously meta-analyzed in the TER, we did not compute a PS, as there was no prior evidence base in the TER for comparison. These associations are narratively described instead.

## Results

### Action 1: Published meta-analyses and pooled analyses

A total of 487 publications (366 meta-analyses, 121 pooled analyses), reporting 2,676 estimates (results) for 1,092 exposure-outcome pairs across 18 exposure groups and 22 cancer sites were included. The average number of primary studies was 8 in the meta-analyses (range: 2-84) and 5 (range: 2-21) in the pooled analyses. Minimal overlap was observed among pooled analyses, affecting only three associations reporting different exposure contrasts (high vs. low and dose-response). A total of 76.1% publications included prospective cohort studies, 4.8% case-control studies, and the rest a mixture of study designs. Most results pertained to body measurements (13%), dietary and lifestyle patterns (12%), and dietary/supplemental micronutrients (10%). Most results were reported for colorectal (17.8%), breast (13.7%) and stomach (8.8%) cancers. Overall, 32% of the results were statistically significant. (**Supplementary Table 2**).

### Action 2: RCTs

Nineteen publications (four protocols and 15 full publications) from nine RCTs were included. Three RCTs (33%) were conducted in females while the rest included both sexes. The number of participants per RCT ranged from 2,157 to 48,835 (average: 12,252). A total of 66 effect estimates (results) were included, with an average of 6 results per publication (range: 1-18). Nine interventions were identified, mainly focusing on vitamin D, calcium, omega-3 supplementations, or combinations of those. Controls comprised of placebo or no intervention groups. Results covered eight cancer sites and cancer precursors, mostly pertaining to colorectal adenomas/polyps (25.8%), followed by breast (22.7%) and prostate (15.2%) cancers. Only three associations were statistically significant showing a protective intervention effect (calcium + vitamin D on skin cancer; omega-3 on prostate cancer; omega-3 + home strength exercise on prostate cancer) (**Supplementary Table 2**).

### Action 3: MR studies

A total of 174 publications, reporting 2,437 effect estimates (results) for 1,634 exposure-outcome pairs across 18 exposure groups and 22 cancer sites were included. The average number of results per publication was 14 (range: 1-304), mostly focusing on micronutrients (31%), body measurements (30.1%) and coffee and tea (7.4%). Most results pertained to colorectal (14.7%), lung (11%), and breast (10.3%) cancers. Overall, 21% of the results were statistically significant in the main MR analysis (**Supplementary Table 2**).

### Action 4: Large cohort studies

A total of 391 publications, reporting 5,924 estimates (results) across 18 exposure groups and 24 cancer sites were included. Most results came from the UK Biobank (44.4%), the European Prospective Investigation into Cancer and Nutrition (EPIC) (19.1%) and NIH-AARP Diet and Health Study (NIH-AARP) (9.1%). Most results focused on body measurements (35.4%), dietary and lifestyle patterns (11.7%) and dietary/supplemental micronutrients (8.8%). Most results pertained to colorectal (17%), breast (9.1%), lymphatic and hematopoietic (8.6%), and prostate (7.8%) cancers. About 31% of the results were statistically significant (**Supplementary Table 2**).

### Action 5: Statistical assessment of the TER findings

Statistical evaluation of the TER results showed that in the majority of associations the expected number of future studies needed to change the previous inference from significant to non-significant (FSN statistic) and from non-significant to significant (CP statistic) greatly exceeded the number of studies in the TER meta-analyses (**Supplementary Figures 10-11**), reflecting general robustness of the TER results. In only two associations (serum alpha-carotene and lung cancer, and toenail selenium and stomach cancer), the FSN was lower than the number of studies in the TER (**Supplementary Table 1**). Conversely, for 36 associations CP indicated a lower expected number of studies compared to the TER, most of which were marginally non-significant in the TER. These included: bladder cancer and vegetable consumption; breast cancer and alcohol (only among former/never users of hormone therapy) and coffee; colorectal cancer and red and processed meat (in females); rectal cancer and poultry, red and processed meat, serum/plasma folate, and total alcoholic drinks; colon cancer and glycemic load, rectal cancer and height (in males); kidney cancer and citrus fruits and processed meat, lung cancer and eggs, height, occupational physical activity, and vegetables (in males); esophageal cancer and citrus fruit and recreational physical activity; esophageal adenocarcinoma and height; pre- and post-menopausal ovarian cancer and body mass index (BMI); pancreatic cancer and total physical activity; prostate cancer and cruciferous vegetables, eggs, serum retinol, serum/plasma/toenail selenium (advanced prostate cancer), whole milk (non-advanced prostate cancer), and wine; melanoma (in males) and BMI and coffee; basal cell carcinoma and decaffeinated coffee; and stomach cancer and eggs, non-fermented soya foods, and processed meat (in females) (**Supplementary Table 1**).

When we compared the TER findings against the new meta-analyses, only for six associations the number of new studies in the meta-analyses exceeded the estimated number of average-sized studies needed to alter previous null meta-analytical estimates from the TER using CP. These associations included: folate and associated compounds and colorectal cancer (17 new studies [excluding the number of studies already in the TER] vs 6 expected to change previous meta-analytic inference); waist circumference and lung cancer (11 new studies vs 3 expected); poultry and breast cancer (10 new studies vs 9 expected); total fat and ovarian cancer (8 new studies vs 5 expected); tea and ovarian cancer (6 new studies vs 4 expected); red meat and kidney cancer (4 new studies vs 3 expected). Using FSN, no new meta-analysis included more studies compared to what was expected to change the significant TER results to null.

### Action 6: Data prioritization summary

A total of 5,152 exposure-cancer associations across 151 exposures and 28 cancer sites were used to compute the PS for exposures included in the TER meta-analyses (**Supplementary Table 2**). We identified 325 exposure-cancer sub-analyses, representing subgroup or population-specific associations related to TER exposures but not previously meta-analyzed. Of these, 114 were from Asian populations, while the remainder primarily focused on subgroups defined by factors such as smoking, diabetes, or obesity (**Supplementary Table 2**).

The median exposure-based PS was 6, ranging from −15 (fruits and vegetables) to 163 (BMI) (**Supplementary Table 2**). Higher scores indicate topics more prominent for prioritization of future SLRs. **Table 2** and **Figure 2** present the top 25 ranked exposures across cancers and within each of the prioritized cancers. Body composition measures ranked highest overall (BMI 1^st^ with PS 163, waist circumference 2^nd^ with PS 110, waist to hip ratio 5^th^ with PS 82, weight gain with PS 54, hip circumference with PS 42, and height with PS 40). Other highly ranked exposures included physical activity (3^rd^ with PS 100) and sedentary behavior (6^th^ with PS 64). The rest of the top-25 ranked exposures for prioritization included vitamins D, C, E, B6, and B12 (ranging from PS 22 for B12 to PS 91 for vitamin D, ranked 4^th^), alcohol (PS 52), tea (PS 36), milk and dairy products (PS 29), micronutrients (iron, dietary fiber, calcium, carotenoids, retinol, and isoflavones, ranging from PS 27 for retinol to PS 38 for iron), potatoes (PS 22), and sugar sweetened beverages (PS 22). Within each exposure and based on the exposure-cancer association PS score, colorectal cancer ranked in the top 5 for 23 of 25 exposures, followed by breast (20/25), lung (16/25), liver (10/25), and endometrial (9/25) cancer. Excluding the 23 associations with strong-convincing evidence grade in the TER only slightly affected the top 25 rankings (± 2 for most exposures) based on the calculated PS score. Height (from 12^th^ to 30^th^) and alcohol (8^th^ to 31^st^) had the larger shifts, replaced by artificial sweetened beverages and zinc in the top 25 list. In addition, waist circumference replaced BMI as the top-ranked exposure (dropped from 1^st^ to 5^th^) (**Supplementary Tables 3-4**). For cancer-specific PS rankings across the 28 sites, colorectal cancer ranked 1^st^, followed by breast, liver, skin, and lymphomas (**Supplementary Table 5**), without any changes in the sensitivity analysis. Cancers of the small intestine and other sites outside the main cancer groups consistently ranked lowest in terms of PS (**Supplementary Tables 5-6**).

**Table 2:**
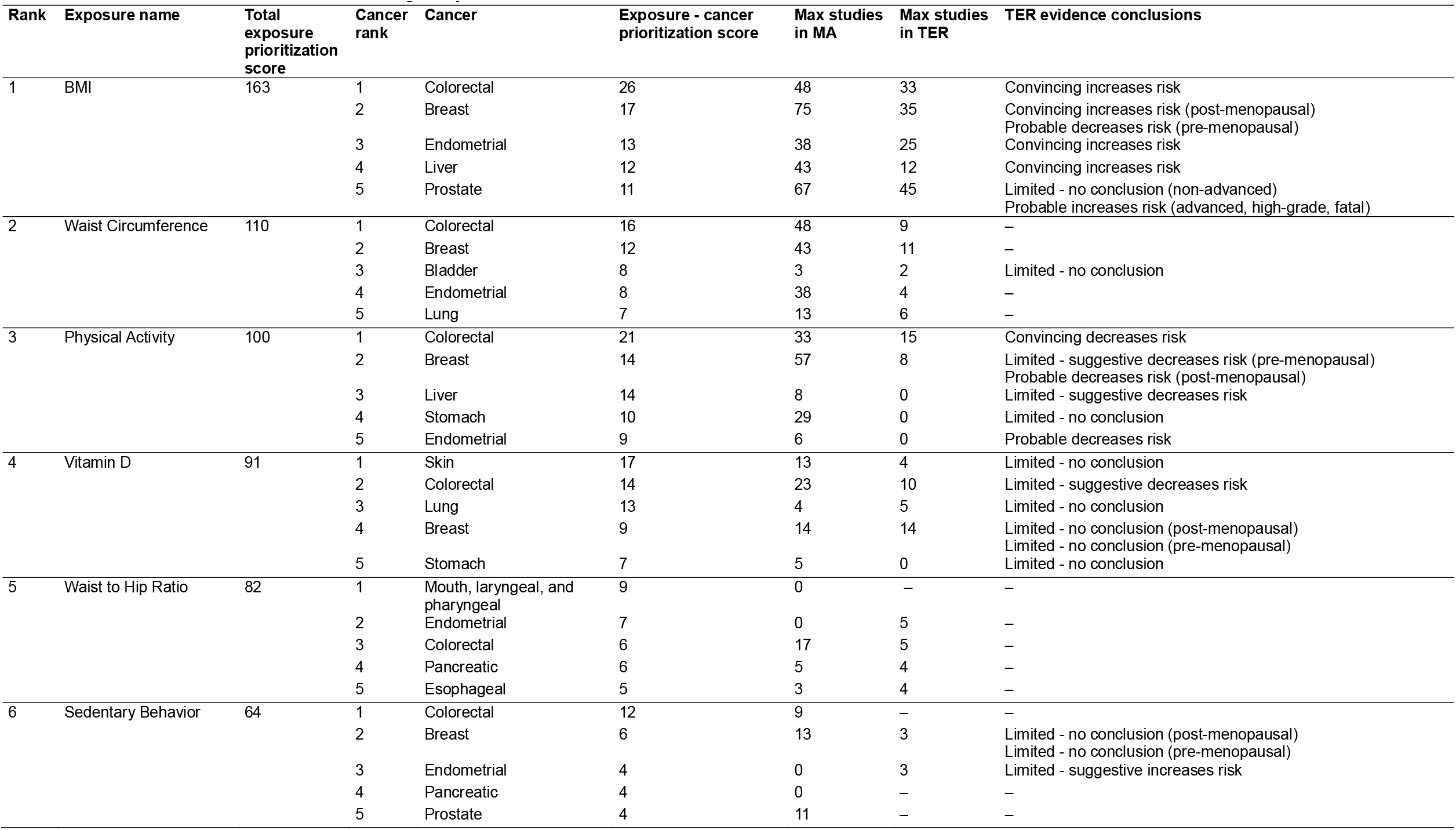
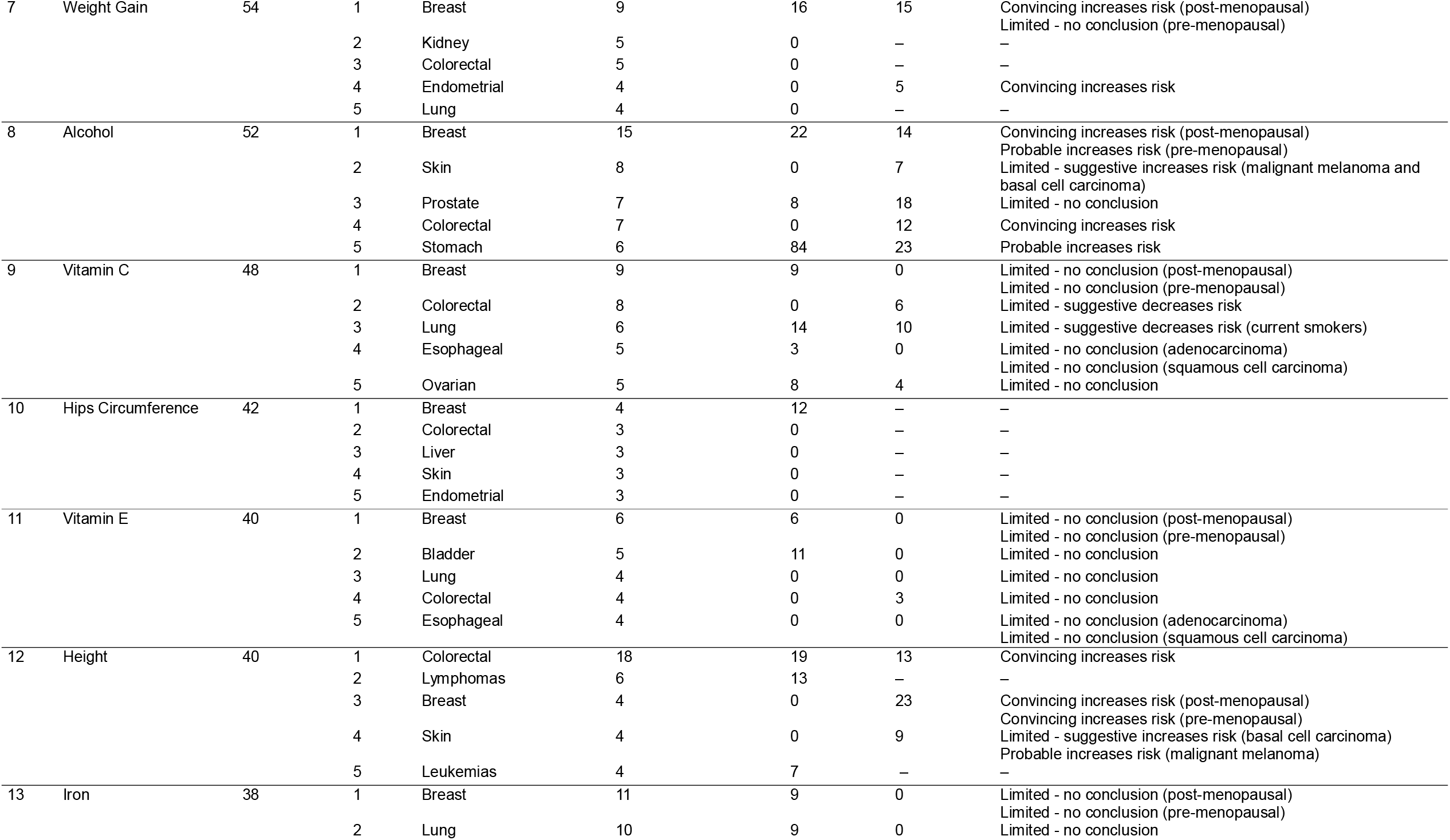
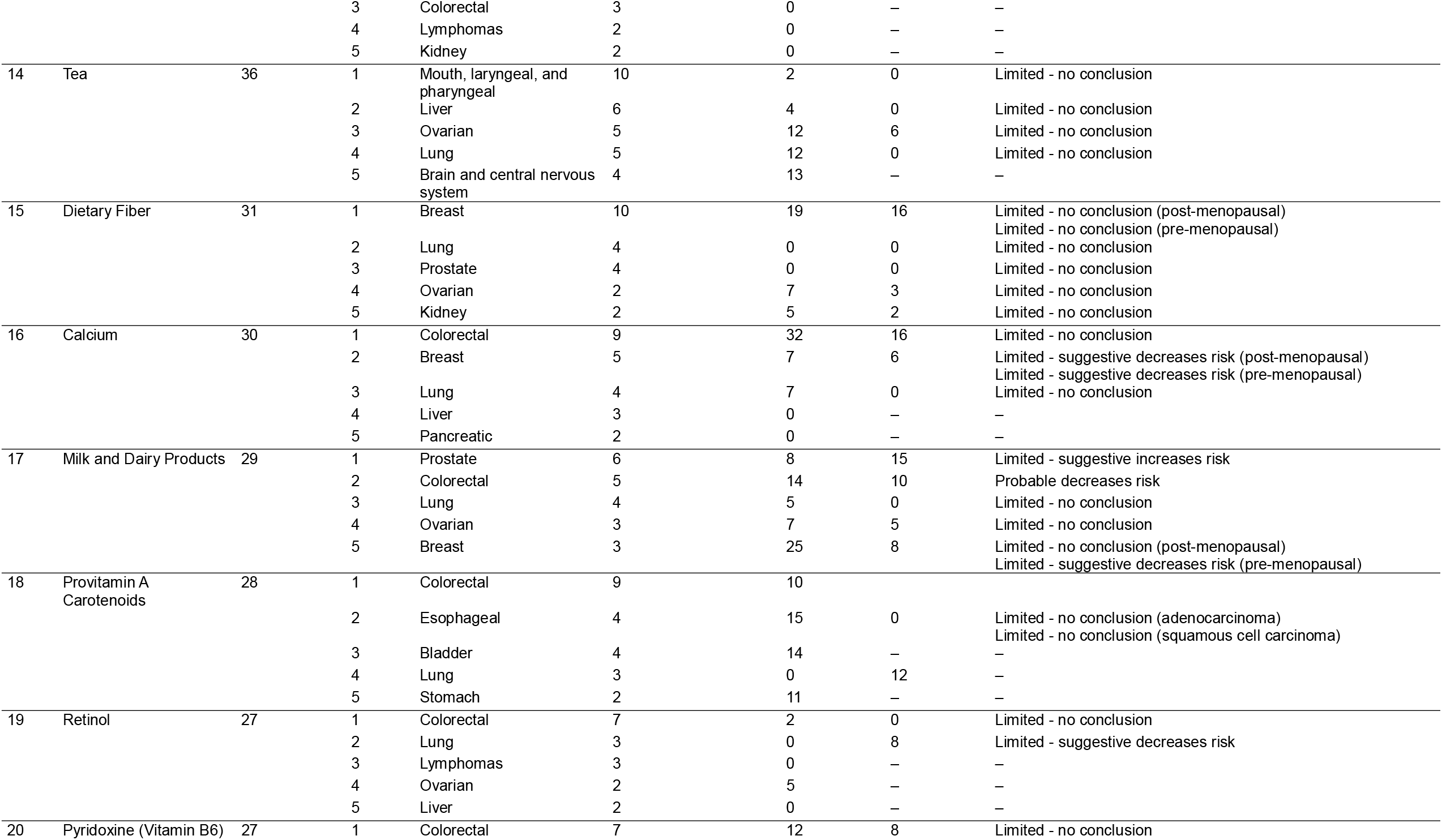
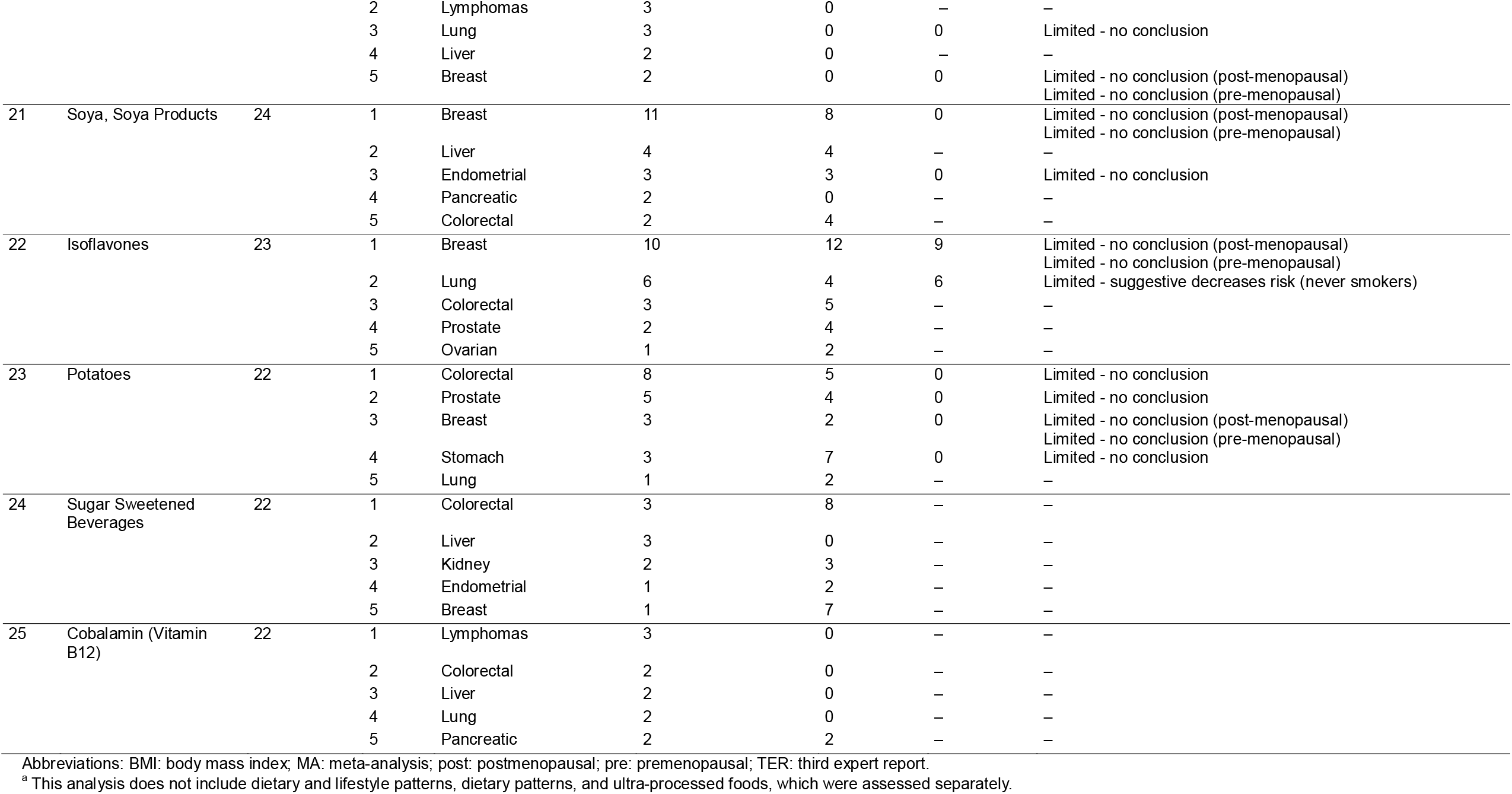
List of the top 25 scoring exposure categories across all cancer outcomes along with the top 5 cancer outcomes per exposure category, followed by the association-specific prioritization score. The final score and the association-specific scores consider all relevant main and subgroup associations ^a^.

**Figure 2:**
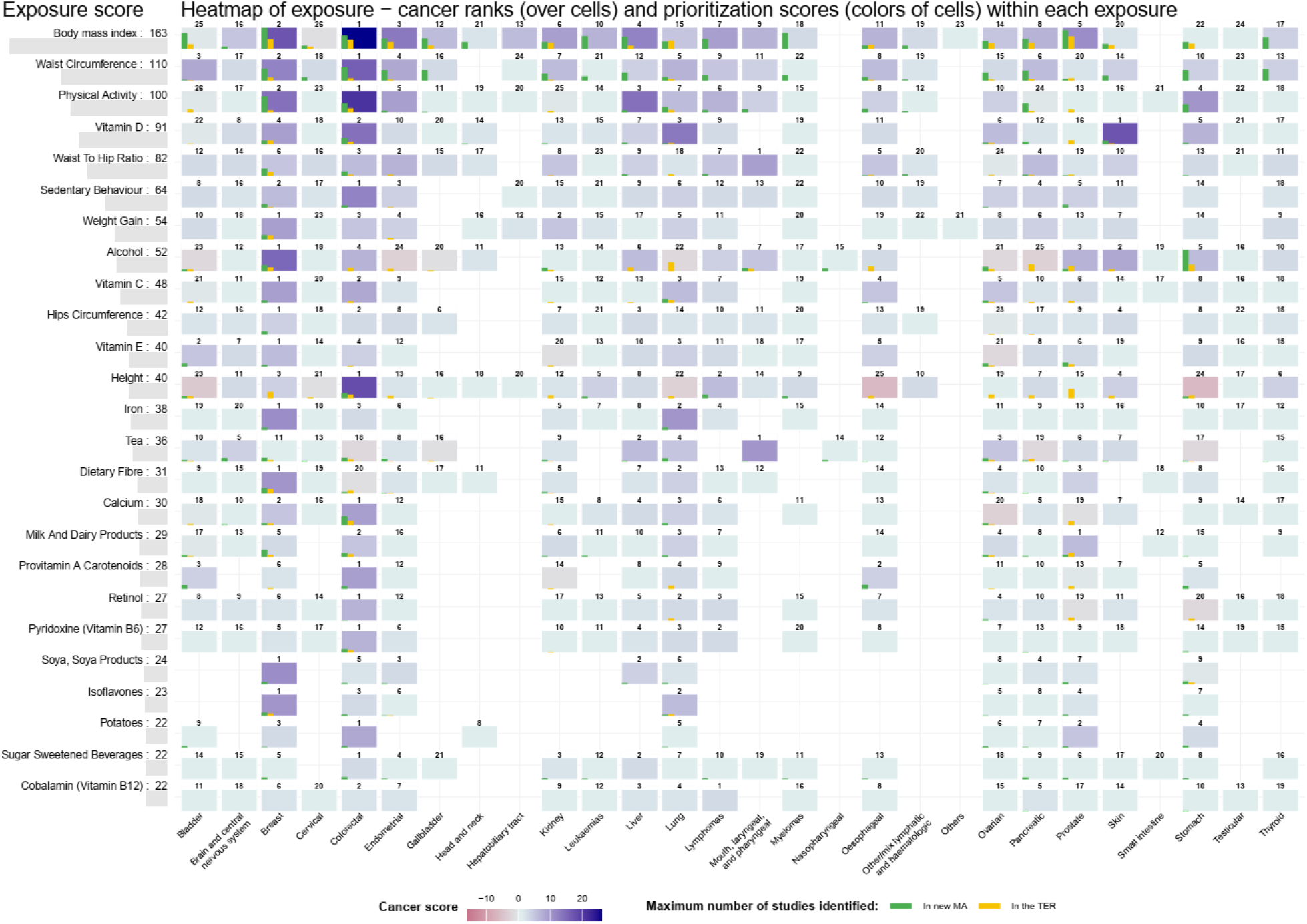
Heatmap depicting the prioritization of exposure-cancer associations based on the exposure-based prioritization score (PS) ^a^. The numbers above the cells represent the ranks of exposure-cancer associations within each exposure, while the color intensity corresponds to the exposure-cancer association PS within each exposure, with higher values (blue color) indicating higher exposure-cancer PS, highlighting areas with the highest potential for future research. ^**a**^ Analyses do not include dietary and lifestyle patterns, dietary patterns, and ultra-processed foods, which were assessed separately.

We identified 1,219 novel associations on exposures not included in the TER meta-analyses, for which PSs were not computed. Of these, 113 were covered in at least two actions or three large cohorts (**Supplementary Table 7**). Five associations were observed across all actions, including nuts with breast, colorectal, lung, and prostate cancers, and low-fat milk with colorectal cancer. Finally, 97 new exposure-cancer associations in meta-analyses of at least three primary studies but not in the TER or other actions were identified, covering 49 novel exposures across 18 cancer sites, such as dietary nitrite and pancreatic, processed vegetables and nasopharyngeal, refined cereals and stomach, fried foods and stomach, carrots and lung, and childhood BMI and colorectal cancers (**Supplementary Table 8**). These associations highlight potential new opportunities to expand the breadth of exposures assessed in the TER.

## Discussion

The present work builds upon the WCRF International’s CUP Global foundation to enable more efficient and timely systematic reviews on the associations between adiposity, diet, physical activity, sedentary behavior and cancer risk. Our results show a substantial accumulation of evidence since the publication of the TER in 2018, with over 1,000 new publications of several study designs, presenting the potential to refine or clarify past conclusions or assess emerging research of novel exposure-cancer associations.

By systematically identifying and prioritizing new and emerging evidence, we examined a large body of literature, including meta-and pooled analyses, RCTs, MR studies, and large cohort studies, to explore the evolving landscape of cancer prevention research. Additionally, using FSN and CP, we evaluated the robustness of the TER findings, and developed a novel scoring algorithm to rank and prioritize exposures based on their potential impact on the evidence base. The utility of data surveillance and prioritization frameworks for updating SLRs has been widely recognized in the literature **(23–25)**. Our framework not only facilitates effective clarification of existing associations, but also enables the identification of critical evidence gaps, and discovery of new research directions. Our findings underscore the significance of targeted prioritization strategies for enhancing and expanding the evidence base on modifiable lifestyle factors in cancer prevention.

The potential to change past conclusions is central to updating SLRs **(25)**. Assessment of the TER results using the CP and FSN statistics offers valuable insights into their robustness and potential future inference shifts. The FSN results suggest that most statistically significant TER associations are highly stable, requiring a substantially large-number of future studies to change their inference. The CP results suggest that the majority of insignificant TER associations are unlikely to change with the addition of future studies with some exceptions, mainly pertaining to associations with sparse evidence in the TER such as folate and colorectal, waist circumference and lung, poultry and breast, total fat and ovarian, tea and ovarian, and red meat and kidney cancers. Overall, these analyses suggest that while the statistically significant associations reported in the TER can be regarded as stable, there is some potential for advancing our understanding of non-significant ones, thus highlighting the importance of targeted efforts to address evidence gaps.

The proposed prioritization framework offers a pragmatic approach for selecting future research directions that could have the highest potential impact on public health policies, ensuring efficient resource allocation. High-ranking exposures such as BMI, waist circumference, and physical activity align with established cancer prevention guidelines **(26–28)**, reinforcing their continued relevance. Colorectal and breast cancers, being two of the most common cancers, achieved high prioritization rankings, reflecting their extensive evidence base. Several of these exposure-cancer associations were already graded as strong-convincing in the TER and are unlikely to be upgraded further. In our prioritisation algorithm, new meta-analyses or pooled analyses contribute to the score primarily when they provide additional evidence for associations that were not statistically significant in the TER. This design ensures that well-established associations do not automatically dominate the priority list, while inconclusive associations or new emerging associations are highlighted for potential future SLRs. Sensitivity analyses excluding associations with prior strong-convincing conclusions showed that our prioritisation remained largely unchanged, with only minor shifts in rank for a few exposures. In addition, the accumulated evidence allows for more in-depth assessment of the associations through subgroup analyses by adiposity, smoking or comorbidity (e.g., diabetes) status. These analyses could refine our understanding of population-specific associations and inform targeted prevention strategies, complementing the current recommendations in alignment with broader public health messages.

Emerging topics such as sugar and artificial sweetened drinks highlight areas that require further investigation. Previously underexplored cancers within the remit of WCRF network such as hematologic neoplasms can now be investigated to address persistent knowledge gaps. The identification of novel exposure-cancer associations not previously covered in the TER, such as allometric body composition indices **(29)**, body fat and other body composition measurements, micronutrients, diet or lifestyle patterns, food mutagens, reflects the dynamic nature of research in the field. Evidence from previously underrepresented populations, notably from Asian cohorts, can provide valuable insights into cancer disparities. These understudied associations along with further gaps in the literature highlight the urgent need for original research in the field, ideally from well-conducted, large-scale cohort studies.

The algorithmic nature of the scoring, while systematic, does not incorporate expert opinions or field-specific insights. Specifically, associations that received a high PS in the present analysis, such as BMI (and obesity in general) with several cancers, may not become immediate priorities for future WCRF International’s CUP Global SLRs due to the relative clarity of the existing public health messages. To tailor the framework’s applicability to WCRF International’s CUP Global priorities, broader considerations were integrated, including institutional objectives, scientific priorities, policy relevance, and feasibility. As a result, high PS-ranked exposures such as sedentary behaviour (ranked 6^th^) and sugar and artificially sweetened beverages (ranked 24^th^), but also lower-ranked exposures such as coffee (ranked 48^th^), were selected for immediate future SLRs. This selection reflects ongoing efforts to address gaps in the evidence and recommendations on modifiable lifestyle factors, including diet, nutrition, anthropometric measures, physical activity, and sedentary behaviour, in relation to cancer risk, as outlined in the recent CUP Global protocol **(30)**.

This framework has several limitations that warrant careful consideration. Despite implementing components of a systematic review across a broad field of study designs, we did not aim for a comprehensive synthesis of the totality of evidence, but rather for an effective assessment of the research landscape. For example, the list of large cohort studies considered was not exhaustive. Although our selection captures a substantial proportion of the available evidence, there is potential for the evidence landscape to shift with the addition of other studies, which could also affect the calculated PS. The scoring algorithm also has certain limitations. It relies on data from diverse study designs, each carrying inherent limitations that are not accounted for in the final score. We aimed to minimize study overlap by selecting a single publication per exposure-cancer association per action. However, overlap across actions cannot be entirely ruled out. A detailed comparison of primary studies in meta-analyses against large cohorts was not conducted, while pooled analyses may share a small number of studies. Nevertheless, this overlap reflects areas with accumulated evidence and active research interest rather than redundancy, supporting the prioritization of these topics for future SLRs. Furthermore, the scoring was primarily based on categorical and linear dose-response analyses reported in the original publications, therefore ignoring non-linear association or more complex analyses like interactions, mediations, or exposure substitution models, which remain sparse in the literature. Finally, emerging research areas in cancer risk also remain underexplored, such as the role of gut microbiota, novel biomarkers, food contaminants, complex modifiable exposures such as diet patterns or ultra processed food consumption. Due to their nature as exposures, these could not be accurately captured in the scores, as the potential of conducting meta-analyses is limited by the inherent heterogeneity of definitions. These domains, while not central to the current prioritization framework, hold significant potential to deepen our understanding of cancer prevention. For instance, ultra-processed foods, although not assigned a PS or rank, were also selected as a topic of interest for future WCRF International’s CUP Global SLRs given their growing relevance and emerging evidence **(30)**.

Our framework, through its facile and effective assessment of the evidence base and the introduction of a novel prioritization score, could have substantial implications for the research community and in public health and policy development. Firstly, it underscores not only the considerable accumulation of new evidence but also the evolving landscape of research on modifiable lifestyle factors, such as adiposity, diet, physical activity, and sedentary behavior in cancer prevention. Second, by identifying the most prominent research fields for future SLRs with the highest potential to shape public health policies, it provides a robust foundation for updating global cancer prevention recommendations.

In conclusion, by integrating the comprehensive evidence synthesis procedures of WCRF International CUP Global with an efficient prioritization framework, we aim to provide a roadmap for future research that will have the potential to better inform public health policies and reduce the global burden of cancer, based on the highest level of evidence.

## Authors’ contribution

Doris S.M. Chan and Konstantinos K. Tsilidis are co-principal investigators of CUP Global at Imperial College London.

Doris S.M. Chan, Georgios Markozannes, and Konstantinos K. Tsilidis developed the prioritization framework and implemented the study.

Katia Balducci, Margarita Cariolou, Sofia Cividini, Ahmad Jayedi, Sayada Zartasha Kazmi, Sonia Kiss, and Rita Vieira did the literature search, study selection, and data extraction.

Sofia Cividini, Doris S.M. Chan, Ahmad Jayedi, Sayada Zartasha Kazmi, Eirini Pagkalidou, and Georgios Markozannes did the data extraction quality checks.

Sofia Cividini, Doris S.M. Chan, Georgios Markozannes, and Konstantinos K. Tsilidis developed the prioritization score.

Ahmad Jayedi and Georgios Markozannes analyzed and interpreted the data.

Dagfinn Aune was a CUP Global Imperial College London team member who revised the manuscript.

Amanda J Cross was a CUP Global advisor at Imperial College London who revised the manuscript.

Marc J. Gunter was the co-lead of the mechanistic data evaluation team at CUP Global who revised the manuscript.

Georgios Markozannes drafted the original manuscript.

All authors reviewed and provided comments on the manuscript.

The work reported in this paper has been performed by the authors, unless clearly specified in the text.

## Acknowledgements

Dr Panagiota Mitrou and Dr Helen Croker, on behalf of the CUP Global Secretariat.

## Data availability statement

Only publicly available data were used in our study. Data sources and handling of these data are described in the materials and methods section. Further details are available from the corresponding author upon request.

## Ethics statement

Ethical approval was not required, since this work was based solely on the analysis of published data without collection of new data or participant involvement.

## References

1. Papadimitriou N, Markozannes G, Kanellopoulou A, Critselis E, Alhardan S, Karafousia V, et al. An umbrella review of the evidence associating diet and cancer risk at 11 anatomical sites. Nat Commun. 2021;12(1):4579.

2. World Cancer Research Fund International. The history of CUP Global. [Accessed: 27 January 2025] https://www.wcrf.org/research-policy/global-cancer-update-programme/history-of-cup-global/ [Available from: https://www.wcrf.org/research-policy/global-cancer-update-programme/history-of-cup-global/.

3. World Cancer Research Fund/American Institute for Cancer Research. Diet, Nutrition, Physical Activity and Cancer: a Global Perspective. Continuous Update Project Expert Report 2018 Available at dietandcancerreportorg.

4. World Cancer Research Fund International. Our Cancer Prevention Recommendations. [Accessed: 27 January 2025] https://www.wcrf.org/preventing-cancer/cancer-prevention/our-cancer-prevention-recommendations/ [Available from: https://www.wcrf.org/preventing-cancer/cancer-prevention/our-cancer-prevention-recommendations/.

5. Abar L, Sobiecki JG, Cariolou M, Nanu N, Vieira AR, Stevens C, et al. Body size and obesity during adulthood, and risk of lympho-haematopoietic cancers: an update of the WCRF-AICR systematic review of published prospective studies. Ann Oncol. 2019;30(4):528–41.

6. Psaltopoulou T, Sergentanis TN, Ntanasis-Stathopoulos I, Tzanninis IG, Riza E, Dimopoulos MA. Anthropometric characteristics, physical activity and risk of hematological malignancies: A systematic review and meta-analysis of cohort studies. International journal of cancer. 2019;145(2):347–59.

7. Teras LR, Patel AV, Carter BD, Rees-Punia E, McCullough ML, Gapstur SM. Anthropometric factors and risk of myeloid leukaemias and myelodysplastic syndromes: a prospective study and meta-analysis. Br J Haematol. 2019;186(2):243–54.

8. Watts EL, Moore SC, Gunter MJ, Chatterjee N. Adiposity and cancer: meta-analysis, mechanisms, and future perspectives. medRxiv : the preprint server for health sciences. 2024.

9. Choi JB, Myong JP, Lee Y, Kim I, Kim JH, Hong SH, et al. Does increased body mass index lead to elevated prostate cancer risk? It depends on waist circumference. BMC cancer. 2020;20(1):589.

10. Choi JB, Lee EJ, Han KD, Hong SH, Ha US. Estimating the impact of body mass index on bladder cancer risk: Stratification by smoking status. Scientific reports. 2018;8(1):947.

11. Choi JB, Kim JH, Hong SH, Han KD, Ha US. Association of body mass index with bladder cancer risk in men depends on abdominal obesity. World journal of urology. 2019;37(11):2393–400.

12. Hwang S, Park YM, Han KD, Yun JS, Ko SH, Ahn YB, et al. Associations of general obesity and central obesity with the risk of hepatocellular carcinoma in a Korean population: A national population-based cohort study. International journal of cancer. 2021;148(5):1144–54.

13. Tran TXM, Kim S, Song H, Ryu S, Chang Y, Park B. Consecutive gain and loss in body weight and waist circumference with risk of subsequent breast cancer in Korean women. International journal of obesity (2005). 2022;46(10):1742–8.

14. Park B. Changes in weight and waist circumference during menopausal transition and postmenopausal breast cancer risk. International journal of cancer. 2022;150(9):1431–8.

15. Elbasheer MMA, Bohrmann B, Chen Y, Lv J, Sun D, Wu X, et al. Reproductive factors and risk of lung cancer among 300,000 Chinese female never-smokers: evidence from the China Kadoorie Biobank study. BMC cancer. 2024;24(1):384.

16. Oze I, Ito H, Koyanagi YN, Abe SK, Rahman MS, Islam MR, et al. Obesity is associated with biliary tract cancer mortality and incidence: A pooled analysis of 21 cohort studies in the Asia Cohort Consortium. International journal of cancer. 2024;154(7):1174–90.

17. Lee S, Jang J, Abe SK, Rahman S, Saito E, Islam R, et al. Association between body mass index and oesophageal cancer mortality: a pooled analysis of prospective cohort studies with >800□000 individuals in the Asia Cohort Consortium. International journal of epidemiology. 2022;51(4):1190–203.

18. Katagiri R, Iwasaki M, Abe SK, Islam MR, Rahman MS, Saito E, et al. Reproductive Factors and Endometrial Cancer Risk Among Women. JAMA network open. 2023;6(9):e2332296.

19. Shin A, Cho S, Jang D, Abe SK, Saito E, Rahman MS, et al. Body Mass Index and Thyroid Cancer Risk: A Pooled Analysis of Half a Million Men and Women in the Asia Cohort Consortium. Thyroid : official journal of the American Thyroid Association. 2022;32(3):306–14.

20. Shin A, Cho S, Abe SK, Islam MR, Rahman MS, Saito E, et al. Association of female reproductive and hormonal factors with gallbladder cancer risk in Asia: A pooled analysis of the Asia Cohort Consortium. International journal of cancer. 2024;155(2):240–50.

21. Roloff V, Higgins JP, Sutton AJ. Planning future studies based on the conditional power of a meta-analysis. Statistics in medicine. 2013;32(1):11–24.

22. Rosenberg MS. The file-drawer problem revisited: a general weighted method for calculating fail-safe numbers in meta-analysis. Evolution; international journal of organic evolution. 2005;59(2):464–8.

23. Ahmadzai N, Newberry SJ, Maglione MA, Tsertsvadze A, Ansari MT, Hempel S, et al. A surveillance system to assess the need for updating systematic reviews. Systematic reviews. 2013;2:104.

24. Shekelle PG, Motala A, Johnsen B. AHRQ Methods for Effective Health Care. Assessment of a Method to Detect Signals for Updating Systematic Reviews. Rockville (MD): Agency for Healthcare Research and Quality (US); 2014.

25. Garner P, Hopewell S, Chandler J, MacLehose H, Schünemann HJ, Akl EA, et al. When and how to update systematic reviews: consensus and checklist. BMJ (Clinical research ed). 2016;354:i3507.

26. World Health Organization. Preventing Cancer. [Accessed: 27 January 2025] https://www.who.int/activities/preventing-cancer [Available from: https://www.who.int/activities/preventing-cancer.

27. Rock CL, Thomson C, Gansler T, Gapstur SM, McCullough ML, Patel AV, et al. American Cancer Society guideline for diet and physical activity for cancer prevention. CA: a cancer journal for clinicians. 2020;70(4):245–71.

28. Schüz J, Espina C, Villain P, Herrero R, Leon ME, Minozzi S, et al. European Code against Cancer 4th Edition: 12 ways to reduce your cancer risk. Cancer epidemiology. 2015;39 Suppl 1:S1–10.

29. Krakauer NY, Krakauer JC. A new body shape index predicts mortality hazard independently of body mass index. PloS one. 2012;7(7):e39504.

30. Global Cancer Update Programme. Protocol for the data collection and systematic literature reviews on the role of diet, sedentary behaviour and the risk of cancer. [Accessed: 27 March 2025] https://osf.io/7utbm/

