## Supplements for "A literature scanning and prioritization framework to guide future systematic reviews for World Cancer Research Fund International’s Global Cancer Update Programme"

**Supplementary material**

### Supplementary Text I – List of cohort studies included in Action 4

1. Cancer Prevention Study II Nutrition Cohort (CPS II Nutrition Cohort)
2. European Prospective Investigation into Cancer and Nutrition (EPIC)
3. Health Professionals Follow-up Study (HPFS)
4. Japan Public Health Center-based Prospective Study (JPHC)
5. Multiethnic Cohort Study (MEC)
6. Netherlands Cohort Study (NLCS)
7. NIH-AARP Diet and Health Study (NIH-AARP)
8. Nurses’ Health Study (NHS)
9. Prostate, Lung, Colorectal and Ovarian Cancer Screening Trial (PLCO)
10. Shanghai Men’s Health Study (SMHS)
11. Shanghai Women’s Health Study (SWHS)
12. UK Biobank
13. Women’s Health Initiative (WHI)

### Supplementary Text II – Search terms for Actions 1-4

a. Simplified PubMed search terms for CUP Global exposures and cancer:

(((("Diet, Food, and Nutrition"[Mesh] OR diet*[tiab] OR nutrition[tiab] OR intake[tiab] OR vitamin[tiab]) OR ("Recreation"[Mesh] OR "Exercise"[Mesh] OR "Healthy Lifestyle"[Mesh] OR "Sedentary Behavior"[Mesh] OR "physical activit*" [tiab] OR "physical inactivity"[tiab] OR "healthy lifestyle"[tiab] OR sedentary[tiab]) OR ("Body Constitution"[Mesh] OR "Body Weight"[Mesh] OR "Body Size"[Mesh] OR "body composition"[tiab] OR weight[tiab] OR height[tiab] OR "body mass index"[tiab] OR "weight loss"[tiab] OR "weight gain"[tiab] OR obesity[tiab] OR adiposity[tiab])) AND ("Neoplasms"[Mesh] OR (cancer*[tiab] OR neoplasia[tiab] OR neoplasm*[tiab] OR tumor*[tiab] OR tumour*[tiab] OR malign*[tiab] OR carcinoma[tiab] OR adenocarcinoma[tiab] OR sarcoma[tiab]))) NOT ("animals"[Mesh] NOT "humans"[Mesh]))

Note: To be combined with b, c, e, or f across the Actions.

b. PubMed search terms for systematic reviews, meta-analyses, and pooled analyses in Action 1:

((systematic[sb] OR meta-analysis[pt] OR "systematic review"[tiab] OR "systematic literature review"[tiab] OR metaanalysis[tiab] OR "meta analysis"[tiab] OR metanalyses[tiab] OR "meta analyses"[tiab] OR "pooled analysis"[tiab] OR "pooled analyses"[tiab] OR "pooled data"[tiab] OR "pooling studies"[tiab] OR "pooling project"[tiab]) NOT ("comment"[Publication Type] OR "editorial"[Publication Type]))

c. PubMed search terms for randomised controlled trials in Action 2:

*Modify a to include cancer precursors, such as “adenomatous polyps”[tiab] OR serrated polyps[tiab] OR “cervical intraepithelial neoplasia”[tiab]

(("randomized controlled trial"[tiab] OR "randomised controlled trial"[tiab] OR "intervention”[tiab]) NOT (survivor*[tiab] OR patients[tiab] OR "comment"[Publication Type] OR "editorial"[Publication Type] OR "systematic review"[Publication Type] OR "meta-analysis"[Publication Type]))

d. Search strategy in clinicaltrials.gov/

Intervention/treatment: diet OR physical activity OR exercise OR sedentary OR weight

Then select in the following filters under Focus Your Search:

Study status/Looking for participants: not yet recruiting OR recruiting.

Study status/No longer looking for participants: active, not recruiting OR completed

Study status/Other: enrolling by invitation OR unknown

Eligibility Criteria/Accepts healthy volunteers: Yes

Study Type: Interventional

Study Results: Without results

Study Documents: Study protocols

More Ways to Search/Outcome measure: cancer and cancer precursor

e. Search terms for MR studies in Action 3:

("Mendelian Randomization Analysis"[Mesh] OR "Mendelian Randomization"[tiab] OR "Mendelian Randomisation"[tiab] OR "genetic instrumental variable"[tiab] OR "genetic instrument"[tiab])

f. Search terms for large cohorts or biobank studies in Action 4:

"Cancer Prevention Study"[tiab] OR "European Prospective Investigation into Cancer and Nutrition"[tiab] OR "Health Professionals Follow-up Study"[tiab] OR “HPFS”[tiab] OR "Japan Public Health Center-based"[tiab] OR "Multiethnic Cohort Study"[tiab] OR "Netherlands Cohort Study"[tiab] OR "NIH-AARP"[tiab] OR "Nurses' Health Study"[tiab] OR "Prostate, Lung, Colorectal and Ovarian Cancer Screening Trial"[tiab] OR “PLCO”[tiab] OR "Shanghai Men's Health Study"[tiab] OR "Shanghai Women's Health Study"[tiab] OR "UK Biobank"[tiab] OR "Women’s Health Initiative"[tiab]

### Supplementary Text III – Description of the data management and analysis procedures, and the prioritization algorithm

First, we excluded non-useable data (e.g., data without effect estimates or evaluations of statistical significance) from the data collected for Actions 1-4. Then, we harmonized the remaining data across all actions to ensure consistency and comparability of results. This procedure included harmonizing exposure and outcome terms across actions and further categorization into main exposure groups, based on the pre-established WCRF exposure codes (ref to our previous protocol) and on specific cancer groups (e.g., colon and rectal subsites were grouped into colorectal cancer group).

The data analysis process involved assigning/aligning the specific exposure-cancer association pairs into results line. The primary pair consisted of the main exposure and cancer association that included both sexes (except for the sex-specific cancers) and for the general population (i.e., not specific population subgroups). In non-sex-specific cancers, if sex was not reported it was assumed to be both men and women. If no specific sub-population was defined, it was assumed that the results referred to all participants. Analyses on cancer incidence, combined cancer incidence & mortality, and cancer mortality were performed separately. -. When the outcome was not explicitly reported, it was assumed to represent incidence. Specifically, in studies using registry data or linked record sources where the outcome definition was not clearly defined (e.g., reported only as “risk”), the outcome was classified as incidence. Although some of these studies may have captured both incident cases and deaths, such composite ascertainment in initially healthy populations predominantly reflects incident diagnoses and was therefore considered most comparable to incidence. Subgroup analyses by sex, by specific cancer sub-sites and on specific populations (e.g., among Asians, among smokers etc.) were considered as separate analyses across all actions. Subgroup analyses were treated separately to provide additional evidence for targeted recommendations and to allow the prioritization algorithm to detect associations that are potentially stronger, more specific, or particularly relevant for certain populations. For each exposure-cancer association pair, we considered both linear dose-response and high vs low comparisons were recorded across all actions.

**Action 1a – Meta-analyses**

We only considered meta-analyses that included at least two primary studies. When multiple meta-analyses were available for a particular exposure-cancer association pair, we selected only **one line of results** to ensure clarity and to avoid redundancy and potential conflicts in data interpretation. Firstly, we prioritized meta-analyses consisting exclusively of cohort studies. If no such meta-analysis was available, we then considered meta-analyses that included a mix of cohort studies and other study designs (e.g., case-control studies)-. Meta-analyses of only case-control studies were not taken into account. Secondly, among the available meta-analyses, we prioritized those that incorporated the largest number of individual studies, which would indicate the breadth of potential new evidence that can be subsequently incorporated in the upcoming SLRs.

**Action 1b – Pooled analyses**

Pooled analyses may have sufficient statistical power to potentially change the previous meta-analytic estimates or may investigate other emerging research topics of interest that are not yet studied in meta-analyses.

When multiple pooled analyses were available for a particular exposure-cancer association pair, results from all pooled analyses were considered, in contrast to Action 1a where a single meta-analysis was selected. We opted for this approach because considering all pooled analyses provides a more comprehensive understanding of the studied association across different studies. We further kept separately all results from the same pooled analysis if more than one lines of association existed but by utilizing studies of different designs.

The advantage of this approach is that it considers all possible different studies that provide information on the association of interest and helps with the identification of patterns and possible discrepancies of the results across studies. The disadvantage is that the studies may overlap between the pooled analyses.

**Action 2 - RCTs**

Results from RCTs were not incorporated in the prioritization algorithm due to most of them being considered mixed interventions or cancer sites that could not be directly aligned to a specific exposure or cancer site of interest or were not statistically significant. The total number of intervention-cancer pairs identified from RCTs was 47, reported across 66 associations, of which 18 (~30%) were on mixed interventions-. Only 3 associations were statistically significant (Calcium + Vitamin D on skin cancer; Omega-3 on prostate cancer; Omega-3 + simple home strength exercise program on prostate cancer) all showing an inverse direction of the association. Non-statistically significant results were not used to de-prioritize associations due to worries that this could be due to lack of power.

**Action 3 – Mendelian randomization analyses**

MR studies can provide supporting (or opposing) evidence for the associations evaluated by observational and interventional studies. The studies can also provide input on possible mechanistic evidence.

For the purpose of creating the data prioritization score, exposure-cancer association lines were not considered when the direction or the significance of the association was not reported. We performed further data harmonization on the population ancestries of the exposure and cancer GWAS to ensure comparability with the other actions. Specifically, we considered European, Asian and Mixed ancestries. Participant characteristics were defined as follows: "All" when both the exposure and cancer population ancestries were either European or not reported (assumed as European), "Asian" when both exposure and cancer population ancestries were Asian, and "Mixed ancestry" when either the exposure population ancestry was Asian and the outcome population ancestry either European or not reported, or vice versa. Reporting as “All” was deliberate so that it could be directly aligned to the overall results from other actions. Results considering Asian ancestry were aligned with the respective results in other actions. The few (only 15) associations categorized as mixed ancestry were not directly aligned to other actions and were excluded. Of those only two were statistically significant (BMI and colorectal cancer and vitamin D and esophageal cancer).

When multiple MR analyses were available for a particular exposure-cancer association pair, we selected only one line of results. We selected the MR analysis that included the highest number of cancer cases, assuming that this MR analysis had the most power to observe an association, to avoid redundancy and potential conflicts in data interpretation that could be due to lack of sufficient power in the non-selected MR results. We further captured information on whether the main MR result was supported by established sensitivity analyses, but this additional information was not utilized for the calculation of the data prioritization score.

**Action 4 – Large cohorts**

Large= cohorts may have sufficient statistical power to potentially change the previous meta-analytic estimates. They may also investigate other emerging research topics of interest. Tracking these publications complements the searches of published meta-analyses that could be outdated.

For the purposes of creating the data prioritization score, we only considered results from large cohorts when both the significance (statistically significant, non-statistically significant) and the direction (positive, inverse) were reported for either a high vs low comparison or a linear dose-response analysis. Exposure outcome pairs that reported non-linear dose-response associations were not considered in the creation of the data prioritization score. For each result line, we kept all reported results across all cohorts and counted the number of statistically significant positive associations, the number of non-statistically significant associations, and the number of statistically significant inverse associations.

**Action 5 – Statistical assessment of the TER findings**

For the statistically significant associations reported in the TER, we computed the fail-safe number (FSN). FSN is a statistic used to estimate the number of additional "null" studies (studies with non-statistically significant findings of average weight similar to those included in the observed meta-analysis) needed to nullify the overall association observed in a previous meta-analysis. This summary estimate is taken directly from the TER. FSN is useful in evaluating the stability of the meta-analytic estimate. It provides insight into the robustness of the meta-analytic findings and indicates whether further investigations are likely to alter the existing conclusions. A high estimated FSN suggests that the current findings are stable and unlikely to be overturned by additional studies, whereas a low estimated FSN indicates that only a few additional null studies could change the significance of the results.

For the non-statistically significant associations reported in the TER, we computed the conditional power (CP) statistic. CP can help predict the potential impact of future studies on the meta-analytic estimate and significance, by assessing whether additional studies are likely to reinforce or alter the current conclusions. CP can be used to determine whether additional studies are worth pursuing. For the purposes of the data prioritization, CP was translated into the estimated number of future studies (of average weight as those included in the observed meta-analysis) required to achieve a CP of at least 80% to detect a nominally statistically significant estimate equal to the observed meta-analytic summary estimate, assuming that the heterogeneity of the updated meta-analysis did not change. The summary estimate is taken directly from the TER. A high estimated N based on a a-priori defined 80% CP indicates a high number of future studies (of similar size and magnitude of the estimate as in the TER) in order to yield significant results, suggesting that further research might not substantially change the current understanding or that larger studies may be needed. In contrast, a low estimated N suggests that only few future studies (of similar size and magnitude of the estimate as in the TER) is needed in order to achieve statistical significance, indicating that additional research is likely to affect the current findings.

The estimated number of studies based on FSN and CP, compared against the number of studies identified by this prioritization exercise, provides an indication of whether an updated meta-analysis could be capable of changing the current inference.

**Action 6: Prioritization score**

The proposed prioritization framework can help guide future SLRs by focusing efforts on the most prominent areas, accounting for the volume, statistical significance and direction of the estimate of new studies after the 2018 the TER. By assigning points based on these criteria, the framework facilitates strategic decision-making about which areas to prioritize for future research.

The following algorithm was used for the creation of a prioritization score that aimed to systematically rank exposure-cancer association pairs based on their results across the multiple actions, comparing them against the results from Action 5 (TER) in terms of concordance of the direction of the estimate and the statistical significance, while also taking into account the estimated number of studies expected to change the inference of the TER result. This score integrates findings from Action 1a: meta-analyses, Action 1b: pooled analyses, Action 3: MR studies, and Action 4: large cohort studies. Non-statistically significant results from Action 2: RCTs and Action 3: MR studies were not used to de-prioritize associations due to worries that this could be due to lack of power. The following describes how points were awarded or subtracted in the calculation of the prioritization score.

**Expected and observed number of new studies**

For each association line with results, we calculated the maximum observed new study counts from Action 1a (meta-analyses). For this comparison, we did not differentiate whether the new meta-analyses reported a categorical high vs low analysis or a linear dose-response analysis, assuming that all the new studies can be utilized in an upcoming SLR. For the purposes of comparing the observed and the expected number of studies from CP, we considered the highest number of observed studies from the two analysis types (categorical high vs low, linear dose-response).

We subsequently compared this number against the expected new study counts from Action 5 (TER) as estimated by the CP statistic, which estimated how many new studies (of average size as those already included in the meta-analysis) are expected to provide sufficient power to shift a non-significant summary estimate to significant. To create the score for this comparison, if the observed number of new studies was equal or larger than the expected number of new studies as estimated by CP, +1 prioritization point was awarded. If the observed number of new studies was much lower than the expected number of new studies (less than 1/10^th^) -1 point was subtracted from the specific association line. No points were given or subtracted from the rest of the association lines or results.

**Note**: Originally, we also planned to incorporate this scoring algorithm for statistically significant associations by utilizing the FSN statistic, but later it was decided that this would not meaningfully contribute to the overall score (as it would only serve to deprioritize significant associations reported in smaller meta-analyses which were more likely to have lower evidence grading and thus higher likelihood of an upgraded evidence grading) and therefore FSN was not incorporated in the final scoring. Instead, we reported separately in the description of Action 5 results a list of associations for which the estimated number of new studies was less than the maximum observed number of new studies in the meta-analyses.

**Evaluation of concordance of results between published meta-analyses and the TER**

For each association line with results, we compared the direction and significance of the results of the published new meta-analyses against those reported in the TER. We prioritized comparing results reported using similar contrasts in the two actions (categorical vs categorical, linear dose-response vs linear dose-response). In case this was not possible (e.g., no meta-analysis reported a linear dose-response association), we also compared the categorical results of the meta-analyses against the linear dose-response results from the TER.

To create the score for this comparison, scenarios for awarding and subtracting points were defined based on the level of concordance between the results in the TER and the new meta-analyses.

If both the meta-analysis and the TER reported statistically significant results in the same direction (positive or negative), +1 point was awarded. If the meta-analysis reported statistically significant results but the TER did not, and the direction of the estimate was concordant, +1 point was also awarded. These associations were assumed to have the capacity of being strengthened, as the results were concordant between the Actions.

If the TER reported significant results but the new meta-analysis did not and the direction of the estimate was concordant, +0.5 points were awarded. Similarly, if both the TER and the meta-analysis reported non-significant results but the direction was concordant, +0.5 points were awarded. Despite the concordance of the direction of the associations, these associations were not deemed at a higher level of priority as it was assumed that the new studies may not be able to strengthen the observed association in the TER with the same level of robustness.

If the direction of estimates between the new meta-analysis and the TER was discordant with the new meta-analytical estimate being statistically significant, a point was neither added nor subtracted. The rationale for this decision is that even though the evidence in the TER and in the new meta-analysis is in the opposite direction, these associations should not be considered as neither a higher priority nor a lower priority.

If the TER reported significant results but the new meta-analysis did not report significant results and the directions were discordant, -1 point was subtracted. If both associations in the TER and new meta-analysis were not statistically significant, -1 point was also subtracted, as it was assumed that the new analyses may not be sufficiently powered to provide meaningful changes in the inference. Associations with discordant results are not excluded but are deprioritized and will not be evaluated first. The PS ensures that associations with the strongest and most consistent evidence are assessed first, while keeping discordant associations available for future evaluation.

In the rare cases where results were available from both categorical vs categorical and linear dose-response vs linear dose-response comparisons, an overall concordance score was calculated by selecting the highest concordance score among the different analyses, reflecting the strongest agreement observed.

**Evaluation of concordance of results between published pooled analyses and the TER**

A similar approach to the calculation of the scores for the meta-analyses was used for the calculation of the prioritization scores of the pooled analyses, with the difference that instead of selecting only the largest meta-analysis for this specific comparison we utilized the results of all different pooled analyses, as described in the relevant section above, assuming that these included different primary studies and therefore provided supplementary information to the others. Comparisons of similar contrasts (categorical vs categorical, linear dose-response vs linear dose-response) were prioritized, otherwise we utilized any available data (e.g., categorical results of the pooled analyses against the linear dose-response results from the TER).

Points were awarded or subtracted based on the agreement of the TER results with the results from the new pooled studies. For example, if 4 pooled analyses reported results on the same association line, a maximum of 4 points could be attributed or subtracted based on the concordance of their results against those from the TER. This was done to emphasize widespread agreement across multiple studies.

**Evaluation of concordance of results between published MR analyses and the TER**

A +1 point was awarded when both the TER and MR studies reported statistically significant results in the same direction. In all the other cases, no points were awarded or subtracted.

**Evaluation of concordance of results between published large cohort studies and the TER**

The results from large cohort studies were assessed as those from pooled analyses, meaning that each study that reported results for a specific association line was separately counted. Comparisons of similar contrasts (categorical vs categorical, linear dose-response vs linear dose-response) were prioritized, otherwise we utilized any available data (e.g., categorical results of the large cohort studies against the linear dose-response results from the TER). Points were awarded or subtracted based on the agreement of the TER results with the results from the multiple cohort studies. For example, if 4 cohort analyses reported results on the same association line a maximum of 4 points could be attributed or subtracted based on the concordance of their results against those from the TER. This was done to emphasize widespread agreement across multiple studies.

**Note**: Due to the sheer volume of results reported in the large cohort studies, the direction of the estimate for non-statistically significant association was not extracted. These associations did not attribute nor subtract prioritization points in case of discordance of significance against the results from the TER.

**Incorporating further evidence for exposures previously assessed in the TER**

The prioritization score aimed to provide an exposure-based ranking system. However, exposures previously assessed in the TER for specific cancer sites may also have sufficient evidence for new cancer sites and/or for new sub-analyses that had to be taken into account in the scoring. To incorporate such evidence on specific subgroups of interest (specific populations or sex-specific analyses when relevant), cancer sites or subsites, and results type (incidence, mortality) that were not previously included in the TER, the prioritization algorithm was not meaningful, since there was a “gold standard” (TER) to serve as a benchmark for comparing the results of the new studies. Therefore, we opted for attributing 3 points for each new association line that pertained to an exposure of interest that was previously assessed in the TER (even for another cancer site) and the number of new studies was sufficient for conducting a new meta-analysis (3 or more studies in total across actions 1a: meta-analyses, 1b: pooled studies, and 4: large cohorts), otherwise each new line of results was given 0.5 prioritization points to indicate that some new evidence exists.

**Final prioritization score**

A total prioritization score was calculated at exposure level. Within each exposure, we computed the sum of all concordance scores from the various Actions for each cancer site and also incorporated points from new sites or subgroups, as described above. Finally, the exposure-cancer scores are summed for each exposure to create a total exposure-specific score.

This score represents the overall alignment and support for each association across multiple analytical methods. Association lines showing strong concordance across all actions received higher scores, indicating higher priorities for a potential SLR.

As sensitivity analyses, we: (1) recalculated the total exposure-specific score by excluding from the scores the associations that had previously received a “strong-convincing” evidence grade in the TER, which were assumed of lower priority for a new SLR, and (2) we calculated a cancer-specific prioritization score (instead of an exposure specific one) by summing all exposure-cancer scores across cancer sites (including and excluding associations that had previously received a strong-convincing evidence grade in the TER).

### Supplementary Table 1: List of exposure-cancer associations from the Third Expert Report (TER) for which the expected number of new studies to change inference as estimated by the fail-safe number and conditional power statistics was lower than the observed number of included studies in the TER meta-analysis.

| **Exposure** | **Contrast** | **Cancer** | **TER findings** | | | **Expected number of new studies to change inference** |
| --- | --- | --- | --- | --- | --- | --- |
|  |  |  | **Number of studies** | **RR (95% CI)** | **I^2^** |  |
| **Statistically significant associations** | | | | | | **Fail-safe number** |
| Serum alpha-carotene | per 10 μg/100 mL | Lung | 5 | 0.44 (0.31, 0.64) | 0.0 | 4 |
| Toenail selenium | High vs. low | Stomach | 2 | 0.58 (0.36, 0.92) | 0.0 | 1 |
| **Non-statistically significant associations** | | | | | | **Conditional power number** |
| Vegetables | per 1 serving/day | Bladder | 9 | 0.97 (0.94, 1.00) | 14.0 | 7 |
| Alcohol | per 10 g/day | Breast (former/never use of MHT) | 3 | 1.12 (1.00, 1.25) | 17.2 | 2 |
| Coffee | per 1 cup/day | Breast | 14 | 0.99 (0.98, 1.00) | 9.0 | 12 |
| Total fat | per 20 g/day | Breast (post-menopausal) | 8 | 1.08 (1.00, 1.17) | 65.9 | 6 |
| Glycemic load | per 50 units/day | Colon | 10 | 0.97 (0.94, 1.00) | 0.0 | 6 |
| Height | per 5 cm | Rectal (male) | 10 | 1.02 (1.00, 1.05) | 43.0 | 9 |
| Poultry | per 100 g/day | Rectal | 6 | 0.85 (0.72, 1.01) | 0.0 | 4 |
| Red and processed meat | per 100 g/day | Colorectal (female) | 8 | 1.13 (0.99, 1.29) | 48.9 | 6 |
| Red and processed meat | per 100 g/day | Rectal | 6 | 1.17 (0.99, 1.39) | 48.1 | 5 |
| Serum/plasma folate | per 2 ng/ml | Rectal | 4 | 0.96 (0.91, 1.00) | 0.0 | 3 |
| Total alcoholic drinks | per 1 drink/day | Rectal | 6 | 1.08 (1.00, 1.17) | 62.5 | 4 |
| Total folate | per 100 mcg/day | Colorectal | 7 | 0.99 (0.98, 1.00) | 0.0 | 6 |
| Citrus fruits | per 50 g/day | Kidney | 4 | 0.98 (0.97, 1.00) | 4.0 | 3 |
| Processed meat | per 50 g/day | Kidney | 3 | 1.05 (1.00, 1.10) | 0.0 | 2 |
| Eggs | per 20 g/day | Lung | 5 | 0.96 (0.92, 1.00) | 0.0 | 4 |
| Height | per 5 cm | Lung | 8 | 1.01 (1.00, 1.02) | 69.1 | 7 |
| Occupational physical activity | High vs. low | Lung | 7 | 1.12 (0.99, 1.28) | 0.0 | 6 |
| Vegetables | per 100 g/day | Lung (male) | 9 | 0.94 (0.88, 1.01) | 49.6 | 7 |
| Citrus fruit | per 100 g/day | Esophageal | 6 | 0.86 (0.74, 1.01) | 0.0 | 4 |
| Height | per 5 cm | Esophageal (adenocarcinoma) | 3 | 0.92 (0.85, 1.00) | 0.0 | 2 |
| Recreational physical activity | High vs. low | Esophageal | 4 | 0.85 (0.72, 1.01) | 0.0 | 3 |
| BMI | per 5 kg/m^2^ | Ovarian (postmenopausal) | 11 | 1.04 (1.00, 1.09) | 47.1 | 8 |
| BMI | per 5 kg/m^2^ | Ovarian (premenopausal) | 6 | 1.12 (1.00, 1.26) | 79.8 | 5 |
| Total physical activity | High vs. low | Pancreatic | 5 | 0.74 (0.55, 1.00) | 47.1 | 3 |
| Cruciferous vegetables | per 50 g/day | Prostate | 7 | 0.96 (0.92, 1.00) | 4.8 | 6 |
| Eggs | per 20 g/day | Prostate (mortality) | 4 | 1.20 (1.00, 1.43) | 41.0 | 3 |
| Serum retinol | per 10 mcg/100ml | Prostate | 11 | 1.01 (1.00, 1.02) | 12.9 | 8 |
| Serum/ plasma/ toenail selenium | per 10mcg/l | Prostate (advanced) | 5 | 0.94 (0.89, 1.00) | 0.0 | 2 |
| Whole milk | per 200 g/day | Prostate (non-advanced) | 4 | 0.94 (0.88, 1.00) | 26.6 | 3 |
| Wines | per 1 drink/day | Prostate | 10 | 1.02 (1.00, 1.04) | 0.0 | 6 |
| BMI | 5 kg/m² | Melanoma (male) | 9 | 1.09 (0.99, 1.19) | 59.2 | 7 |
| Coffee | per 1 cup/day | Melanoma | 7 | 0.96 (0.92, 1.00) | 44.3 | 4 |
| Decaffeinated coffee | per 1 cup/day | Basal cell carcinoma | 3 | 1.02 (1.00, 1.03) | 0.0 | 2 |
| Eggs | per 1 time/w | Stomach | 5 | 1.03 (1.00, 1.06) | 0.0 | 3 |
| Non-fermented soya foods | High vs. low | Stomach | 6 | 0.81 (0.65, 1.00) | 29.2 | 4 |
| Processed meat | per 50 g/day | stomach (female) | 6 | 1.40 (1.00, 1.97) | 0.0 | 3 |

Abbreviations: BMI: body mass index; CI: confidence interval; MHT: menopause hormone therapy; RR: relative risk.

### Supplementary Table 4: Change in exposure ranking based on the prioritization score in a sensitivity analysis excluding exposure-cancer associations with a strong or convincing evidence grade from the **Third Expert Report (TER)**.

| **Main prioritization score** | | **Sensitivity analysis prioritization score** | | **Change in rank** |
| --- | --- | --- | --- | --- |
| **Rank** | **Exposure group** | **Rank** | **Exposure group** |  |
| 1 | Body mass index | 1 | Waist circumference | +1 |
| 2 | Waist circumference | 2 | Physical activity | +1 |
| 3 | Physical activity | 3 | Vitamin D | +1 |
| 4 | Vitamin D | 4 | Waist to hip ratio | +1 |
| 5 | Waist to hip ratio | 5 | Body mass index | -4 |
| 6 | Sedentary behavior | 6 | Sedentary behavior | – |
| 7 | Weight gain | 7 | Vitamin C | +2 |
| 8 | Alcohol | 8 | Weight gain | -1 |
| 9 | Vitamin C | 9 | Hips circumference | 1 |
| 10 | Hips circumference | 10 | Vitamin E | 1 |
| 11 | Vitamin E | 11 | Iron | 2 |
| 12 | Height | 12 | Tea | 2 |
| 13 | Iron | 13 | Dietary fiber | 2 |
| 14 | Tea | 14 | Calcium | 2 |
| 15 | Dietary fiber | 15 | Milk and dairy products | 2 |
| 16 | Calcium | 16 | Provitamin A carotenoids | 2 |
| 17 | Milk and dairy products | 17 | Retinol | 2 |
| 18 | Provitamin A carotenoids | 18 | Pyridoxine (Vitamin B6) | 2 |
| 19 | Retinol | 19 | Soya, soya products | 2 |
| 20 | Pyridoxine (Vitamin B6) | 20 | Isoflavones | 2 |
| 21 | Soya, soya products | 21 | Potatoes | 2 |
| 22 | Isoflavones | 22 | Sugar sweetened beverages | 2 |
| 23 | Potatoes | 23 | Cobalamin (Vitamin B12) | 2 |
| 24 | Sugar sweetened beverages | 24 | Artificial sweetened beverages | 2 |
| 25 | Cobalamin (Vitamin B12) | 25 | Zinc | 2 |
| 26 | Artificial sweetened beverages | 30 | Height | -18 |
| 27 | Zinc | 31 | Alcohol | -23 |

### Supplementary Table 6: Change in cancer ranking based on the prioritization score in a sensitivity analysis excluding exposure-cancer associations with a strong or convincing evidence grade from the **Third Expert Report (TER)**.

| **Main prioritization score** | | **Sensitivity analysis prioritization score** | | **Change in rank** |
| --- | --- | --- | --- | --- |
| **Rank** | **Cancer** | **Rank** | **Cancer** |  |
| 1 | Colorectal | 1 | Colorectal | – |
| 2 | Breast | 2 | Breast | – |
| 3 | Liver | 3 | Liver | – |
| 4 | Skin | 4 | Skin | – |
| 5 | Lymphomas | 5 | Lymphomas | – |
| 6 | Lung | 6 | Lung | – |
| 7 | Endometrial | 7 | Ovarian | +1 |
| 8 | Ovarian | 8 | Endometrial | -1 |
| 9 | Mouth, laryngeal, and pharyngeal | 9 | Prostate | +1 |
| 10 | Prostate | 10 | Mouth, laryngeal, and pharyngeal | -1 |
| 11 | Esophageal | 11 | Brain and central nervous system | +1 |
| 12 | Brain and central nervous system | 12 | Esophageal | -1 |
| 13 | Kidney | 13 | Thyroid | +1 |
| 14 | Thyroid | 14 | Bladder | +1 |
| 15 | Bladder | 15 | Leukemias | +1 |
| 16 | Leukemias | 16 | Kidney | -3 |
| 17 | Pancreatic | 17 | Myelomas | +1 |
| 18 | Myelomas | 18 | Pancreatic | -1 |
| 19 | Other/mix lymphatic and hematologic | 19 | Other/mix lymphatic and hematologic | – |
| 20 | Stomach | 20 | Stomach | – |
| 21 | Hepatobiliary tract cancer | 21 | Hepatobiliary tract cancer | – |
| 22 | Head and neck | 22 | Head and neck | – |
| 23 | Gallbladder | 23 | Gallbladder | – |
| 24 | Cervical | 24 | Cervical | – |
| 25 | Testicular | 25 | Testicular | – |
| 26 | Nasopharyngeal | 26 | Nasopharyngeal | – |
| 27 | Small intestine | 27 | Small intestine | – |
| 28 | Others | 28 | Others | – |

### **Supplementary Table 7: Novel exposure-cancer associations pertaining to exposures that were not meta-analyzed in the Third Expert Report (TER) that were identified in at least 2 different actions or in at least 3 different large cohorts.**

| **Exposure group** | **Cancer** | **Meta-analysis** | | | **Pooled analyses** | | **Large cohorts** | | **MR analysis** |
| --- | --- | --- | --- | --- | --- | --- | --- | --- | --- |
|  |  | **Number of studies** | **Statistical significance** | **Direction of estimate** | **Number of studies** | **Study code** | **Number of studies** | **Study name** |  |
| Gluten | Colorectal |  |  |  | 1 | P8 | 2 | CPSII, UKB |  |
| Gluten | Pancreatic |  |  |  | 1 | P8 | 1 | UKB |  |
| Maize (corn) | Liver |  |  |  | 1 | P6 | 1 | NIH-AARP |  |
| Bread | Colorectal |  |  |  |  |  | 2 | EPIC, UKB | Yes |
| Bread | Lung |  |  |  |  |  | 2 | EPIC, UKB | Yes |
| Refined cereals and cereal products | Colorectal |  |  |  |  |  | 3 | CPSII, MEC, UKB |  |
| Refined cereals and cereal products | Liver |  |  |  | 1 | P6 | 1 | WHI |  |
| Mushrooms | Breast | 10 | Sig* | - |  |  | 1 | EPIC |  |
| Mushrooms | Colorectal | 2 | NS* | - |  |  | 1 | EPIC |  |
| Mushrooms | Prostate | 2 | NS* | - | 1 | P5 |  |  |  |
| Blueberries, strawberries and other berries | Bladder | 2 | NS* | - | 1 | P1 |  |  |  |
| Peanut butter | Colorectal | 2 | NS* | + |  |  | 1 | NLCS |  |
| Peanut butter | Lung | 2 | NS* | - |  |  | 1 | NLCS |  |
| Peanut butter | Ovarian | 2 | NS* | - |  |  | 1 | NLCS |  |
| Nuts | Brain and central nervous system | 2 | NS* | + | 1 | P4 |  |  |  |
| Nuts | Breast | 6 | NS* | - | 1 | P8 | 1 | EPIC | Yes |
| Nuts | Colorectal | 9 | Sig | - | 1 | P8 | 3 | EPIC, MEC, NLCS | Yes |
| Nuts | Endometrial | 2 | NS* | - |  |  | 1 | NLCS | Yes |
| Nuts | Liver | 3 | NS* | - | 1 | P6 |  |  |  |
| Nuts | Lung | 5 | Sig* | - | 1 | P8 | 2 | EPIC, NLCS | Yes |
| Nuts | Esophageal | 5 | NS | - |  |  |  |  | Yes |
| Nuts | Ovarian | 3 | NS* | - |  |  | 1 | NLCS | Yes |
| Nuts | Pancreatic | 5 | Sig | - |  |  | 1 | EPIC |  |
| Nuts | Prostate | 6 | NS* | + | 1 | P8 | 1 | NLCS | Yes |
| White meat | Liver | 6 | NS* | - | 1 | P6 |  |  |  |
| White meat | Stomach | 4 | NS | - | 1 | P11 |  |  |  |
| Lamb | Colorectal |  |  |  |  |  | 2 | EPIC, UKB | Yes |
| Fatty fish | Colorectal |  |  |  |  |  | 2 | EPIC, UKB | Yes |
| White fish, lean fish | Colorectal |  |  |  |  |  | 2 | EPIC, UKB | Yes |
| Butter | Bladder | 5 | NS* | o | 1 | P1 | 1 | PLCO |  |
| Butter | Liver |  |  |  | 1 | P6 | 1 | WHI |  |
| Butter | Lung | 2 | NS* | - |  |  | 1 | EPIC |  |
| Olive oil | Breast | 3 | NS* | - | 1 | P7 |  |  | Yes |
| High-fat dairy | Colorectal | 3 | NS* | - |  |  | 1 | NHS |  |
| High-fat dairy | Liver |  |  |  | 1 | P6 | 1 | WHI |  |
| Low-fat milk | Breast | 6 | NS | + | 1 | P8 |  |  |  |
| Low-fat milk | Colorectal | 2 | Sig* | - | 1 | P8 | 1 | UKB | Yes |
| Low-fat milk | Ovarian | 7 | NS* | + | 1 | P8 |  |  |  |
| Low-fat dairy | Colorectal | 3 | NS* | - |  |  | 1 | NHS |  |
| Low-fat dairy | Liver |  |  |  | 1 | P6 | 1 | WHI |  |
| Cakes, biscuits and pastry | Colorectal |  |  |  |  |  | 2 | EPIC, UKB | Yes |
| Confectionery | Colorectal |  |  |  |  |  | 3 | CPSII, EPIC, UKB | Yes |
| Chocolate, candy bars | Colorectal |  |  |  |  |  | 3 | EPIC, UKB, WHI |  |
| Citrus fruit juice | Skin |  |  |  |  |  | 3 | EPIC, NIH-AARP, WHI |  |
| Orange / grapefruit juice | Liver |  |  |  | 1 | P9 | 1 | NIH-AARP |  |
| Vegetable juices | Liver |  |  |  | 1 | P9 | 1 | NIH-AARP |  |
| Caffeinated coffee | Brain and central nervous system |  |  |  | 1 | P8 | 1 | UKB |  |
| Caffeinated coffee | Colorectal | 3 | NS* | - |  |  | 2 | CPSII, UKB |  |
| Caffeinated coffee | Ovarian | 4 | NS* | + |  |  |  |  | Yes |
| Cadmium | Breast | 17 | Sig* | + | 1 | P2 |  |  |  |
| DiMelQx | Pancreatic | 4 | Sig* | + |  |  | 1 | MEC |  |
| MelQx | Pancreatic | 4 | Sig* | + |  |  | 1 | MEC |  |
| PhIP | Pancreatic | 4 | Sig* | + |  |  | 1 | MEC |  |
| Ratio polyunsaturated/saturated fat | Liver | 2 | Sig | - | 1 | P6 |  |  |  |
| N-3 fatty acids | Breast | 11 | Sig* | - |  |  |  |  | Yes |
| N-3 fatty acids | Colorectal | 19 | Sig* | - |  |  | 1 | UKB | Yes |
| N-3 fatty acids | Endometrial | 12 | NS* | + |  |  |  |  | Yes |
| N-3 fatty acids | Liver | 3 | NS* | - | 1 | P6 |  |  |  |
| N-3 fatty acids | Ovarian |  |  |  |  |  | 2 | EPIC, NHS | Yes |
| N-3 fatty acids | Prostate | 3 | NS* | + |  |  |  |  | Yes |
| N-6 fatty acids | Breast | 9 | NS* | + |  |  | 1 | CPSII | Yes |
| N-6 fatty acids | Colorectal | 13 | NS* | + |  |  | 1 | UKB | Yes |
| N-6 fatty acids | Liver | 2 | NS | - | 1 | P6 |  |  |  |
| Protein | Breast |  |  |  |  |  | 3 | EPIC, UKB, WHI | Yes |
| Plant protein | Prostate | 4 | NS | + |  |  | 2 | EPIC, NIH-AARP |  |
| Animal protein | Colorectal |  |  |  |  |  | 3 | EPIC, UKB, NIH-AARP |  |
| Animal protein | Prostate | 5 | NS | o |  |  | 1 | EPIC |  |
| Dairy protein | Prostate | 3 | Sig | + |  |  | 2 | EPIC, UKB |  |
| Amino acids | Colorectal |  |  |  | 1 | P8 | 2 | EPIC, UKB |  |
| Sodium | Lung |  |  |  | 1 | P10 | 2 | PLCO, WHI |  |
| Non-haem iron | Lung | 3 | NS* | + |  |  | 1 | EPIC |  |
| Magnesium | Colorectal |  |  |  |  |  | 2 | EPIC, UKB | Yes |
| Magnesium | Lung | 5 | NS | - |  |  | 2 | EPIC, PLCO | Yes |
| Potassium | Lung |  |  |  | 1 | P10 | 3 | EPIC, PLCO, WHI |  |
| Polyphenols | Stomach | 7 | NS* | - | 1 | P11 |  |  |  |
| Anthocyanidins | Stomach | 2 | NS* | - | 1 | P11 |  |  |  |
| Flavonoids | Ovarian | 6 | Sig* | - |  |  | 1 | EPIC |  |
| Other bioactive compounds | Breast | 3 | NS* | + |  |  | 2 | EPIC, NHS |  |
| Cardiorespiratory fitness | Colorectal |  |  |  |  |  | 2 | NIH-AARP, UKB | Yes |
| BMI middle adulthood | Pancreatic |  |  |  |  |  | 2 | NIH-AARP, PLCO | Yes |
| Childhood BMI | Ovarian | 5 | Sig* | + |  |  |  |  | Yes |
| Obesity | Breast |  |  |  | 1 | P7 | 1 | UKB | Yes |
| Obesity | Colorectal | 10 | Sig* | + |  |  | 1 | UKB | Yes |
| Obesity | Esophageal | 3 | NS* | + |  |  | 1 | UKB | Yes |
| Obesity | Ovarian |  |  |  |  |  | 1 | UKB | Yes |
| Obesity | Pancreatic | 5 | NS* | + |  |  | 2 | NIH-AARP, UKB | Yes |
| Body fat percentage | Breast |  |  |  |  |  | 2 | UKB, WHI | Yes |
| Body fat percentage | Prostate | 2 | NS | + |  |  | 1 | UKB | Yes |
| Body fat | Breast |  |  |  | 1 | P7 | 2 | UKB, WHI | Yes |
| Body fat | Lung |  |  |  | 1 | P6 | 1 | UKB | Yes |
| Body fat | Ovarian |  |  |  |  |  | 2 | NIH-AARP, UKB | Yes |
| Body fat | Prostate |  |  |  |  |  | 2 | HPFS, UKB | Yes |
| Weight change | Breast |  |  |  | 1 | P7 | 1 | EPIC |  |
| Weight change | Colorectal |  |  |  | 1 | P6 | 1 | EPIC |  |
| Weight change | Kidney | 2 | Sig* | + | 1 | P6 | 1 | EPIC |  |
| Weight loss | Bladder |  |  |  | 1 | P6 | 1 | EPIC |  |
| Weight loss | Brain and central nervous system |  |  |  | 1 | P6 | 1 | EPIC |  |
| Weight loss | Breast |  |  |  | 1 | P3, P6 | 2 | EPIC, WHI |  |
| Weight loss | Colorectal |  |  |  | 1 | P6 | 3 | EPIC, PLCO, WHI |  |
| Weight loss | Hepatobiliary tract cancer |  |  |  | 1 | P6 | 1 | EPIC |  |
| Weight loss | Kidney |  |  |  | 1 | P6 | 2 | EPIC, WHI |  |
| Weight loss | Leukemias |  |  |  | 1 | P6 | 1 | EPIC |  |
| Weight loss | Liver |  |  |  | 1 | P6 | 1 | WHI |  |
| Weight loss | Lung |  |  |  | 1 | P6 | 2 | EPIC, JPHC |  |
| Weight loss | Lymphomas |  |  |  | 1 | P6 | 1 | EPIC |  |
| Weight loss | Myelomas |  |  |  | 1 | P6 | 1 | WHI |  |
| Weight loss | Esophageal |  |  |  | 1 | P6 | 1 | EPIC |  |
| Weight loss | Ovarian |  |  |  | 1 | P6 | 2 | EPIC, WHI |  |
| Weight loss | Pancreatic |  |  |  | 1 | P6 | 2 | EPIC, WHI |  |
| Weight loss | Prostate |  |  |  | 1 | P6 | 1 | EPIC |  |
| Weight loss | Skin |  |  |  | 1 | P6 | 1 | EPIC |  |
| Weight loss | Stomach |  |  |  | 1 | P6 | 1 | EPIC |  |
| Weight loss | Thyroid |  |  |  | 1 | P6 | 2 | EPIC, WHI |  |

Abbreviations: BMI: Body Mass Index; CPS II Nutrition Cohort: Cancer Prevention Study II Nutrition Cohort; EPIC: European Prospective Investigation into Cancer and Nutrition; HPFS: Health Professionals Follow-up Study; JPHC: Japan Public Health Center-based Prospective Study; MEC: Multiethnic Cohort Study; MR: Mendelian randomization; NLCS: Netherlands cohort study; NIH-AARP: NIH-AARP Diet and Health Study; NHS: Nurses’ Health Study; ns: not significant; PLCO: Prostate, Lung, Colorectal and Ovarian Cancer Screening Trial; sig: significant; SMHS: Shanghai Men’s Health Study; SWHS: Shanghai Women’s Health Study; UK Biobank: UK Biobank; WHI: Women’s Health Initiative.

Pooled analysis code: Study name: P1: BLEND; P2: CPSII LifeLink, EPIC-Italy & NSHDS; P3: DCPP; P4: MWS & NIH-AARP & PLCOS; P5: Miyagi & Ohsaki; P6: NHS & HPFS; P7: NHS & NHSII; P8: NHS, NHSII & HPFS; P9: NIH-AARP & PLCO; P10: PLCO & WHI; P11: StoP.

* Denotes that the association is based on categorical contrasts. All exposure contrasts were harmonized in order to reflect higher versus lower levels or doses of the exposure.

### Supplementary Table 8: Novel exposure-cancer associations covered only in action 1 (meta-analysis) of at least 3 primary studies **pertaining to exposures that were not meta-analyzed in the Third Expert Report (TER)**.

| **Exposure group** | **Cancer** | **Number of studies** | **Statistical significance** | **Direction of estimate** |
| --- | --- | --- | --- | --- |
| Rice, pasta, noodles | Stomach | 9 | Sig* | + |
| Refined cereals and cereal products | Breast | 7 | NS | o |
| Refined cereals and cereal products | Stomach | 16 | Sig* | + |
| Root vegetables | Ovarian | 6 | NS* | - |
| Fruiting vegetables | Ovarian | 7 | NS* | - |
| Carrots | Lung | 18 | Sig* | - |
| Onion | Breast | 7 | Sig* | - |
| Dark green vegetables | Bladder | 3 | NS* | - |
| Dark yellow vegetables | Bladder | 4 | NS* | - |
| Dark yellow vegetables | Brain and central nervous system | 4 | Sig* | - |
| Leafy vegetables | Bladder | 5 | NS* | - |
| Leafy vegetables | Ovarian | 3 | NS* | - |
| Mushrooms | Stomach | 5 | NS* | - |
| Bananas | Ovarian | 4 | NS* | + |
| Apples, pears | Ovarian | 5 | NS* | + |
| Soy beans | Ovarian | 3 | NS* | - |
| Peanut butter | Esophageal | 4 | NS* | + |
| Peanut butter | Stomach | 4 | NS* | - |
| Nuts | Leukemias | 3 | NS* | - |
| Nuts | Stomach | 7 | NS* | - |
| White meat | Pancreatic | 9 | Sig | + |
| Butter | Mouth, laryngeal, and pharyngeal | 5 | NS* | - |
| Butter | Ovarian | 5 | NS* | o |
| Margarine | Ovarian | 4 | NS* | + |
| Hard cheese | Ovarian | 3 | NS* | - |
| Cottage cheese | Ovarian | 3 | NS* | - |
| Fermented dairy products | Bladder | 5 | Sig* | - |
| Fermented milk and milk products | Colorectal | 11 | NS* | - |
| Fermented milk and milk products | Lung | 3 | NS* | - |
| Fermented milk and milk products | Pancreatic | 5 | NS* | - |
| Ice cream | Ovarian | 4 | NS* | - |
| Spicy foods | Esophageal | 22 | Sig | + |
| Fruit and vegetable juices | Ovarian | 4 | NS* | o |
| Coffee and tea | Ovarian | 23 | NS* | - |
| Caffeinated coffee | Pancreatic | 3 | NS* | + |
| Cadmium | Ovarian | 3 | NS* | - |
| Nitrate in water | Bladder | 4 | NS* | - |
| Nitrate in water | Breast | 4 | NS* | + |
| Nitrate in water | Colorectal | 9 | Sig* | + |
| Nitrate in water | Pancreatic | 4 | NS* | + |
| Processed vegetable | Nasopharyngeal | 26 | Sig* | + |
| Dietary nitrite | Bladder | 41 | NS* | + |
| Dietary nitrite | Brain and central nervous system | 41 | NS* | + |
| Dietary nitrite | Colorectal | 41 | NS* | + |
| Dietary nitrite | Kidney | 41 | NS* | - |
| Dietary nitrite | Lymphomas | 41 | NS* | + |
| Dietary nitrite | Esophageal | 41 | NS* | + |
| Dietary nitrite | Ovarian | 3 | NS* | + |
| Dietary nitrite | Pancreatic | 41 | Sig* | - |
| Dietary nitrite | Stomach | 41 | NS* | + |
| Fried foods | Stomach | 18 | Sig* | + |
| BaP | Breast | 3 | NS* | - |
| BaP | Lung | 4 | NS* | + |
| BaP | Pancreatic | 4 | NS* | + |
| BaP | Prostate | 7 | NS* | o |
| DiMelQx | Bladder | 4 | NS* | + |
| DiMelQx | Breast | 7 | NS* | - |
| DiMelQx | Colorectal | 7 | NS* | + |
| DiMelQx | Kidney | 3 | NS* | - |
| DiMelQx | Lung | 4 | NS* | + |
| DiMelQx | Esophageal | 3 | NS* | + |
| DiMelQx | Prostate | 11 | NS* | + |
| Heterocyclic amines | Colorectal | 5 | NS* | + |
| Heterocyclic amines | Prostate | 3 | NS* | + |
| MelQx | Bladder | 4 | NS* | + |
| MelQx | Breast | 8 | NS* | + |
| MelQx | Colorectal | 9 | Sig* | + |
| MelQx | Kidney | 3 | NS* | + |
| MelQx | Lung | 5 | Sig* | + |
| MelQx | Esophageal | 3 | NS* | + |
| MelQx | Prostate | 12 | NS* | + |
| MelQx | Stomach | 3 | NS* | - |
| PhIP | Bladder | 4 | Sig* | + |
| PhIP | Breast | 8 | NS* | + |
| PhIP | Colorectal | 9 | NS* | + |
| PhIP | Kidney | 4 | NS* | + |
| PhIP | Lung | 5 | NS* | + |
| PhIP | Esophageal | 3 | NS* | + |
| PhIP | Prostate | 12 | NS* | + |
| PhIP | Stomach | 4 | NS* | + |
| Mutagen index, meat | Colorectal | 4 | NS* | + |
| N-3 fatty acids | Pancreatic | 3 | NS* | - |
| N-6 fatty acids | Endometrial | 12 | NS* | + |
| N-6 fatty acids | Pancreatic | 4 | NS* | - |
| N-6 fatty acids | Prostate | 6 | NS* | - |
| N-6 fatty acids | Skin | 3 | NS* | + |
| Methionine | Breast | 7 | NS* | - |
| Methionine | Pancreatic | 4 | NS* | - |
| Soya protein | Breast | 4 | Sig* | - |
| Vitamin B complex | Ovarian | 5 | NS* | - |
| Caffeine | Ovarian | 6 | NS* | - |
| Anthocyanidins | Breast | 5 | NS* | - |
| Flavonoids | Breast | 8 | NS* | - |
| Flavonoids | Prostate | 3 | NS* | + |
| Moderate-high intensity activity | Stomach | 11 | NS* | - |
| Childhood BMI | Colorectal | 14 | Sig* | + |
| BMI loss | Thyroid | 10 | Sig* | - |

Abbreviations: BMI: Body Mass Index; ns: not significant; sig: significant.

* Denotes that the association is based on categorical contrasts. All exposure contrasts were harmonized in order to reflect higher versus lower levels or doses of the exposure.

### Supplementary Figure 1 The percentage of the results across 18 major exposure groups reported in the published meta-analyses and pooled analyses.

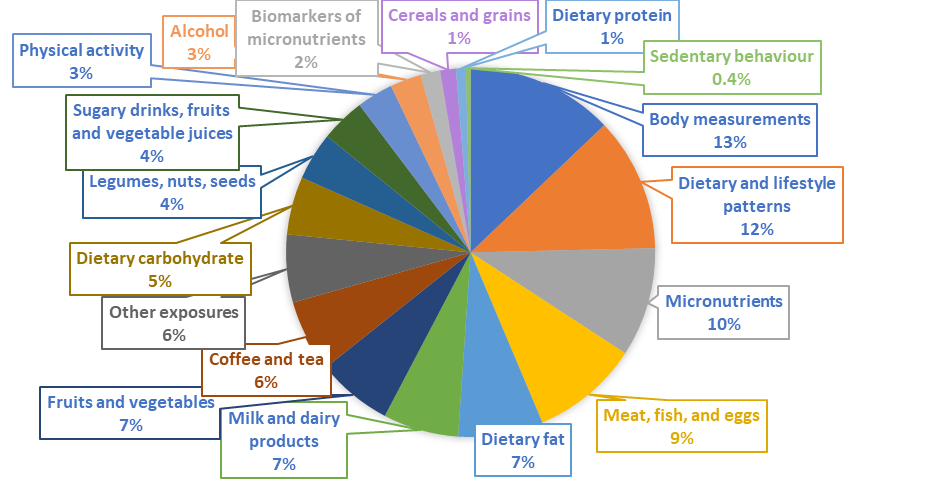

### Supplementary Figure 2 Number of publications and results across 18 major exposure groups reported in the published meta-analyses and pooled analyses.

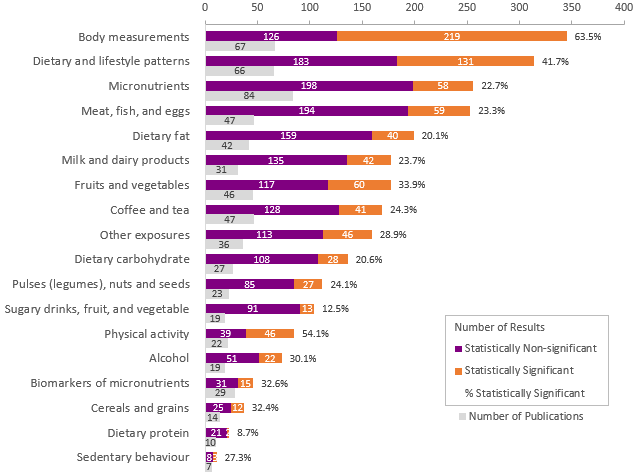

Note: Other exposures group includes exposures such as spicy foods, cakes, biscuits and pastry, confectionery, contaminants in food/water sources, salting and pickling, food processing and preparation, sweeteners, meat mutagens, and energy intake.

### Supplementary Figure 3 Number of publications and results across 24 cancer sites reported in the published meta-analyses and pooled analyses.

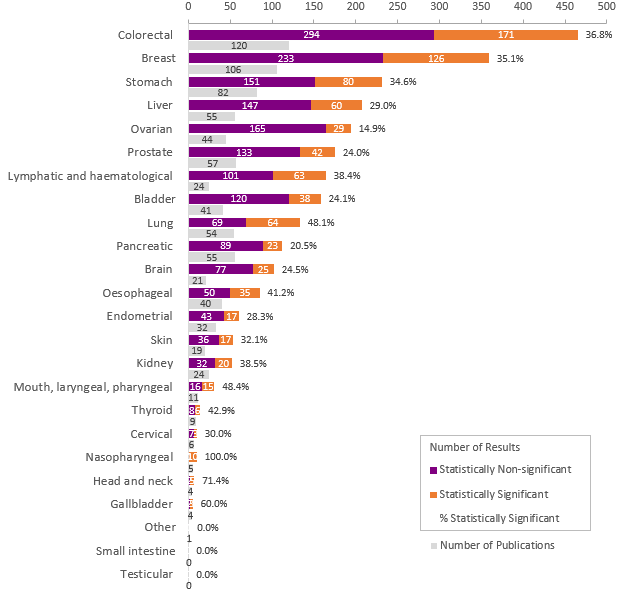

Note: Other cancers include upper aerodigestive cancer. Lymphatic and hematological cancers include leukemia, lymphoma, myeloma, myelodysplastic syndromes, and myeloproliferative neoplasms.

### Supplementary Figure 4 Number of publications and results across 9 dietary and lifestyle interventions.

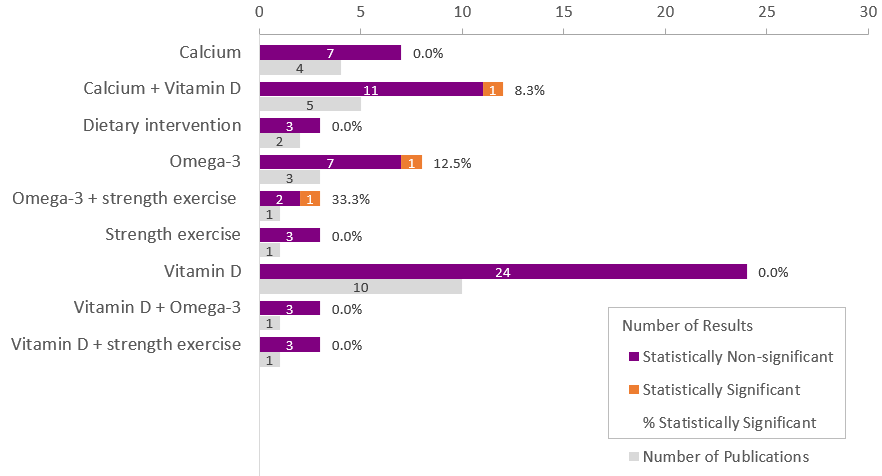

Note: Dietary intervention = low fat diet + ↑ fruits, vegetables, and grains.

### Supplementary Figure 5 Number of publications and results across 8 cancer sites or cancer precursors reported in the randomized controlled trials.

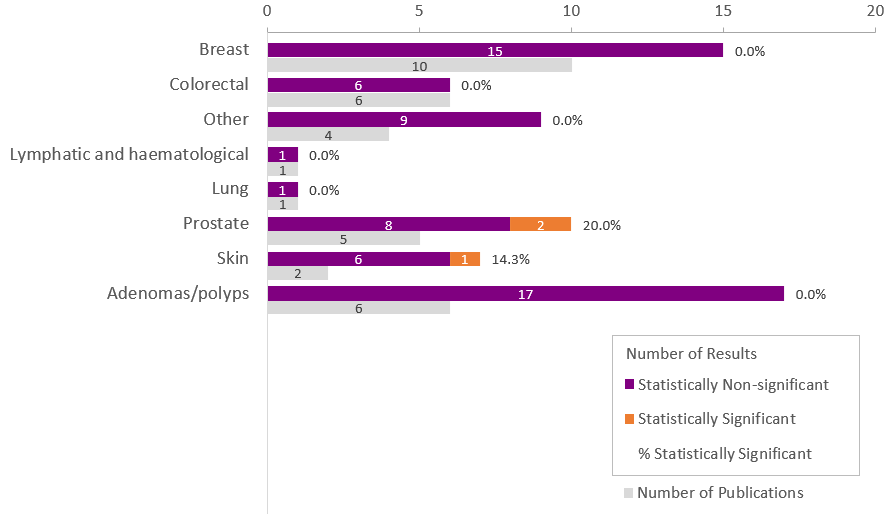

Note: Other cancers include gastrointestinal, orodigestive, and respiratory cancers. Lymphatic and hematological cancers include leukemia, lymphoma, myeloma, myelodysplastic syndromes, and myeloproliferative neoplasms.

### Supplementary Figure 6 Number of publications and results across 18 major exposure groups reported in the Mendelian randomization studies.

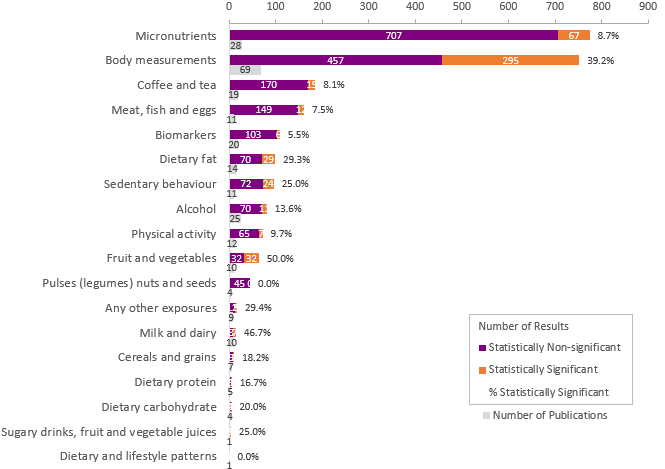

Note: Other exposures include sugar, diet sugar, and crackers/crispbreads with butter/margarine.

### Supplementary Figure 7 Number of publications and results across 24 cancer sites reported in the Mendelian randomization studies.

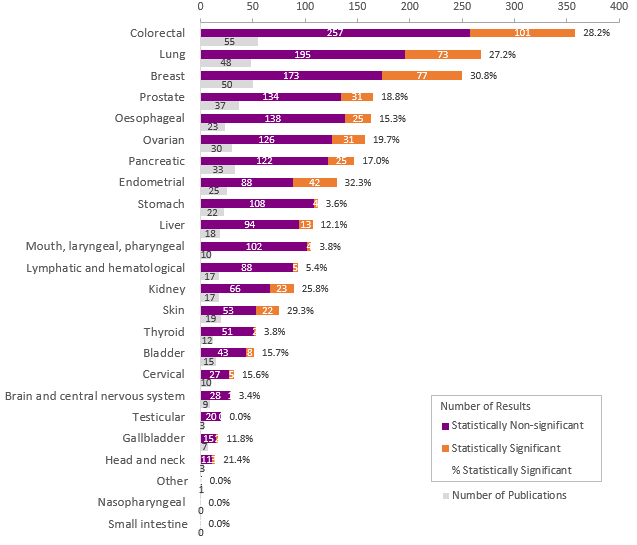

Note: Other cancers include upper aerodigestive and bone cancers. Lymphatic and hematological cancers include leukemia, lymphoma, myeloma, myelodysplastic syndromes, and myeloproliferative neoplasms.

### Supplementary Figure 8 Number of publications and results across 18 major exposure groups reported in the large cohorts.

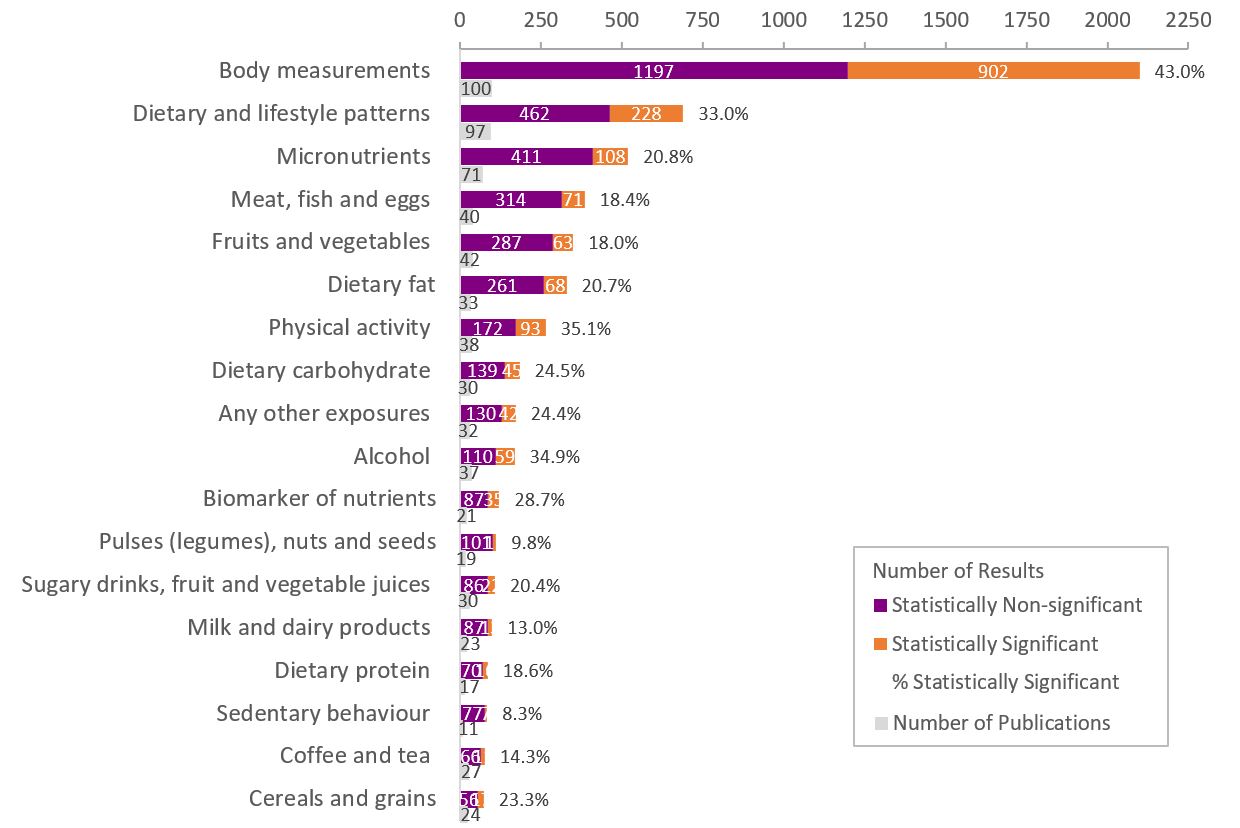

Note: Other exposures include acrylamide, biscuits, cakes, cookies and pastry, confectionery, composite foods, fast foods, heterocyclic amines, lead, cadmium, and other chemicals, energy intake, meat mutagens, mycotoxins, pickles, pizza, resting energy expenditure, sauces, snacks, soups, sugars, vegetable-based dishes or dips, water.

### **Supplementary Figure 9 Number of publications and results across 24 cancer sites reported in the large cohorts.**

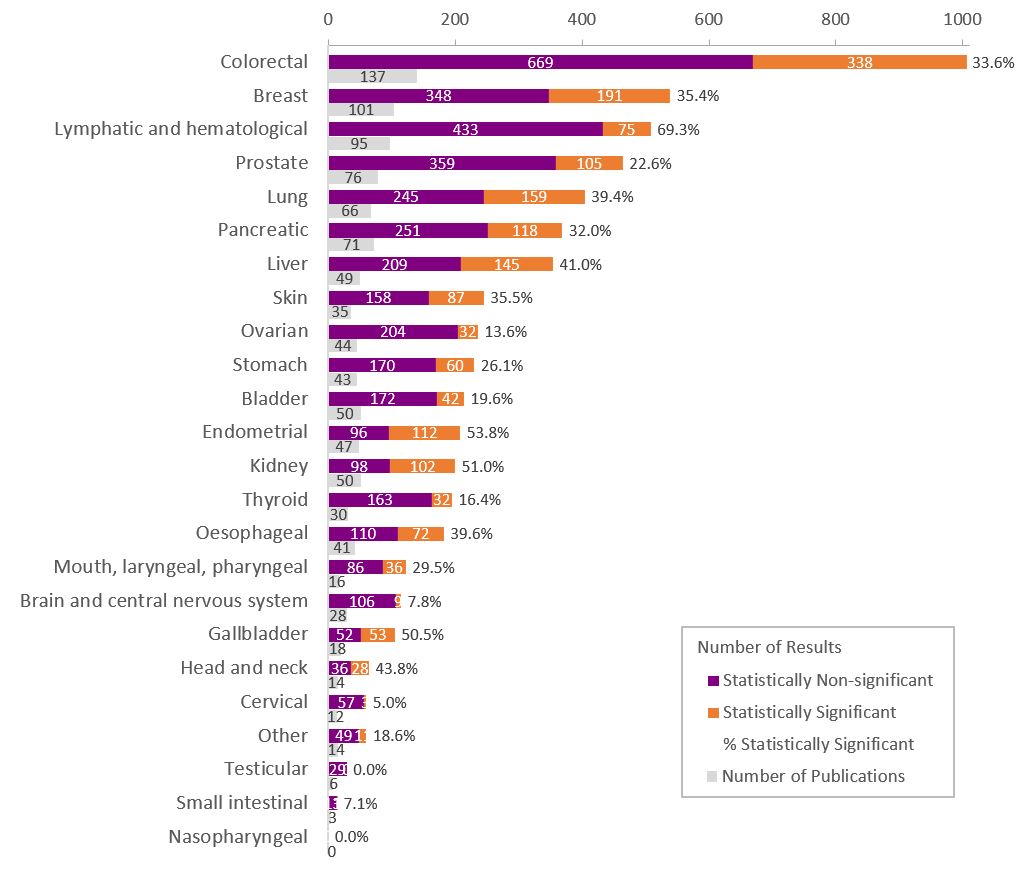

Note: Other cancers include combinations of liver, gallbladder and bile ducts cancers, mesothelioma, upper-aerodigestive cancer, anogenital and anal cancers, and sarcomas. Others/mix lymphatic and hematological cancers include leukemia, lymphoma, myeloma, myelodysplastic syndromes, myeloproliferative neoplasms, and other or mixed lymphatic and hematopoietic tissue cancers.

### **Supplementary Figure 10 Observed number of studies included in the statistically significant meta-analyses from the Third Expert Report (TER) and the estimated number of additional null studies ^a^ needed to nullify the overall association observed in a previous meta-analysis.**

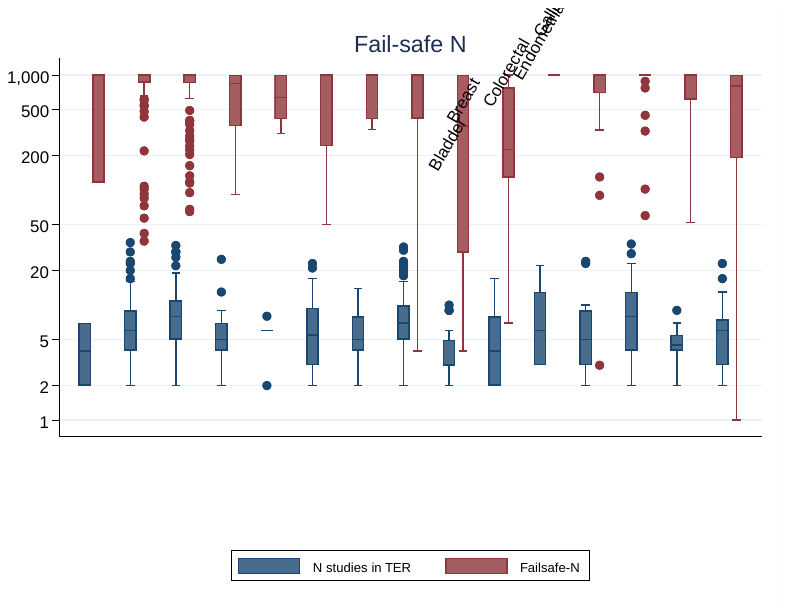

**^a^** Studies with non-statistically significant findings of average weight similar to those included in the observed meta-analysis.

### **Supplementary Figure 11 Observed number of studies included in the non-statistically significant meta-analyses from the Third Expert Report (TER) and the estimated number** of future studies ^a^ required to achieve a conditional power (CP) of at least 80% ^b^.

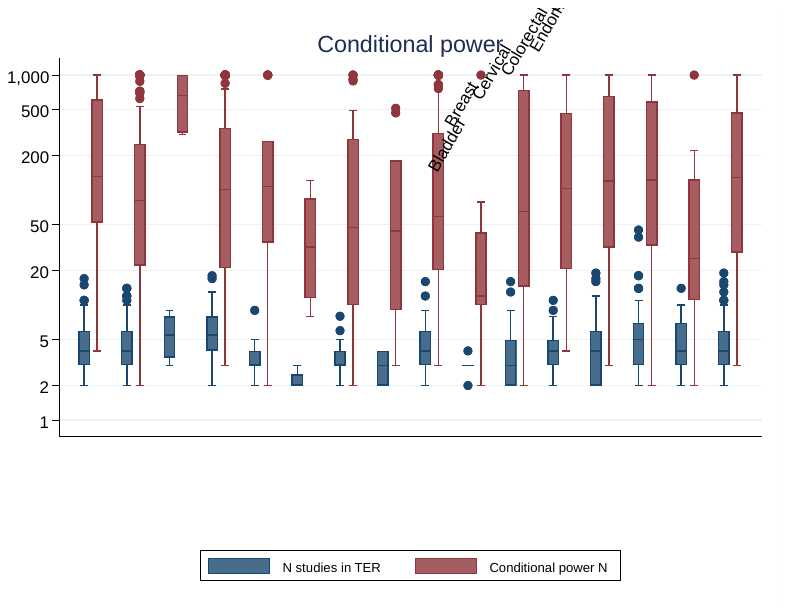

**^a^** Of average weight as those included in the observed meta-analysis

**^b^** To detect a nominally statistically significant estimate equal to the observed meta-analytic summary estimate, assuming that the heterogeneity of the updated meta-analysis did not change.
